# Single nucleus transcriptomics, pharmacokinetics, and pharmacodynamics of combined CDK4/6 and mTOR inhibition in a phase 0/1 trial of recurrent high-grade glioma

**DOI:** 10.1101/2024.06.07.24308439

**Authors:** Kevin C. Johnson, An-Chi Tien, Jun Jiang, James McNamara, Yu-Wei Chang, Chelsea Montgomery, Anita DeSantis, Leonel Elena-Sanchez, Yoko Fujita, Seongho Kim, Avishay Spitzer, Paul Gabriel, William F. Flynn, Elise T. Courtois, Amy Hong, Jocelyn Harmon, Yoshie Umemura, Artak Tovmasyan, Jing Li, Shwetal Mehta, Roel Verhaak, Nader Sanai

## Abstract

Outcomes for adult patients with a high-grade glioma continue to be dismal and new treatment paradigms are urgently needed. To optimize the opportunity for discovery, we performed a phase 0/1 dose-escalation clinical trial that investigated tumor pharmacokinetics, pharmacodynamics, and single nucleus transcriptomics following combined ribociclib (CDK4/6 inhibitor) and everolimus (mTOR inhibitor) treatment in recurrent high-grade glioma. Patients with a recurrent high-grade glioma (n = 24) harboring 1) *CDKN2A*/*B* deletion or *CDK4*/*6* amplification, 2) *PTEN* loss or *PIK3CA* mutations, and 3) wild-type retinoblastoma protein (Rb) were enrolled. Patients received neoadjuvant ribociclib and everolimus treatment and no dose-limiting toxicities were observed. The median unbound ribociclib concentrations in Gadolinium non-enhancing tumor regions were 170 nM (range, 65 – 1770 nM) and 634 nM (range, 68 – 2345 nM) in patients receiving 5 days treatment at the daily dose of 400 and 600 mg, respectively. Unbound everolimus concentrations were below the limit of detection (< 0.1 nM) in both enhancing and non-enhancing tumor regions at all dose levels. We identified a significant decrease in MIB1 positive cells suggesting ribociclib-associated cell cycle inhibition. Single nuclei RNAseq (snRNA) based comparisons of 17 IDH-wild-type on-trial recurrences to 31 IDH-wild-type standard of care treated recurrences data demonstrated a significantly lower fraction of cycling and neural progenitor-like (NPC-like) malignant cell populations. We validated the CDK4/6 inhibitor-directed malignant cell state shifts using three patient-derived cell lines. The presented clinical trial highlights the value of integrating pharmacokinetics, pharmacodynamics, and single nucleus transcriptomics to assess treatment effects in phase 0/1 surgical tissues, including malignant cell state shifts. ClinicalTrials.gov identifier: NCT03834740.

## INTRODUCTION

Standard of care treatment of adult patients with glioblastoma consists of maximal extent of resection followed by concomitant chemoradiation, resulting in a median overall survival of 15 months [1]. The minimal progress that has been achieved in improving treatment outcomes suggests that innovative approaches that diverge from conventional oncological strategies are needed [2]. Here, we present a dual drug phase 0 ‘trigger’ trial that leverages pharmacokinetics (PK), pharmacodynamics (PD), and single-nucleus transcriptome sequencing to enable the discovery of treatment-associated biological signals [3].

Focal gains or amplifications in *CDK4*/*CDK6* and *CDKN2A* homozygous deletion in glioma represent genomic alterations that disrupt normal functioning of the cell cycle [4, 5]. Ribociclib is a highly specific small-molecule inhibitor of *CDK4*/*CDK6* [6], and has been shown to significantly prolong survival outcomes for patients with advanced breast cancer [7]. We previously showed that ribociclib was well tolerated and exhibited good central nervous system (CNS) penetration in a phase 0/1 clinical trial of patients with high-grade glioma [8]. Analysis of tissues from patients who experienced a recurrence while on ribociclib monotherapy treatment detected an increase in the mammalian target of rapamycin (mTOR) pathway activity, suggesting a potential mechanism of resistance. These results prompted initiation of a phase 0/1 trial with ribociclib and a mTOR inhibitor, everolimus, described here. In addition to tumor pharmacokinetics and pharmacodynamics analysis, in-depth molecular analyses of tumor specimens from the phase 0 trial represent an opportunity to quickly and broadly understand mechanisms of action and malignant cellular plasticity in response to investigational drugs.

Single cell and nucleus RNA sequencing studies have previously shown that most glioblastomas consist of malignant cell states that include stem-like populations of neural progenitor cell-like (NPC-like) and oligodendrocyte progenitor-like (OPC-like) cells as well as more differentiated populations of astrocyte-like (AC-like) and mesenchymal-like (MES-like) cells, with subsets of malignant cells in each cell state additionally being characterized by an active cell cycle signature [9–12]. The distributions of malignant cell states have been shown to be influenced by both the microenvironment and genetics, including an association between greater NPC-like signal and *CDK4* amplifications [10]. We have previously shown that direct application of stressors (e.g. hypoxia) results in cell state shifts [12], and that acquired genetic alterations can lead to consistent changes in cell states [13]. The drivers of such transitional shifts may be multifactorial and include therapy or underlying genetic evolution. Profiling human glioma samples that are actively on targeted drugs (e.g., CDK4/6 inhibitors) are needed to understand how these therapies influence tumor cell biology and demonstrate whether it is possible to pharmacologically shift malignant cellular states.

In this current phase 0/1 trial, we investigated the combination of ribociclib (CDK4/CDK6 inhibitor) plus everolimus (mTOR inhibitor) in recurrent high-grade glioma to limit resistance to ribociclib. This study was designed to determine the primary outcome measures of 1) the total and unbound concentrations of both ribociclib and everolimus in contrast enhancing and non-enhancing tumor tissue, 2) identify the molecular effects of both drugs via assessment of proximal pharmacodynamic markers (pRb, pS6 and p4EBP1) and single nuclei transcriptomic analyses, and 3) determine maximum tolerated dosing regimen for the drug combinations. The trial was designed to qualify patients for a phase 1 expansion if a patient’s tumor samples met pharmacokinetic and pharmacodynamic thresholds for both therapies. Overall, our study highlights the importance of integrating pharmacokinetics/pharmacodynamics and post-treatment molecular characterization to assess glioma treatment effects from experimental therapeutics [3].

## RESULTS

### Study population

Patient demographics and clinical characteristics are summarized in **Table 1** (CONSORT diagram; **Extended Data Fig. 1**). Eligibility was determined based on previous clinical records, panel DNA sequencing, and immunohistochemistry of archival tissue from a previous surgery (**Figure 1, Extended Data Fig. 2a**). The inclusion criteria required that all patients had recurrent high-grade glioma with 1) Retinoblastoma protein (Rb) expression or wild-type *RB1* gene, 2) *CDK4/6* amplification or *CDKN2A* deletion and 3) *PTEN* or *PIK3CA* mutations. There were 21 patients with an IDH-wild-type tumor and 3 patients with a high-grade IDH-mutant astrocytoma enrolled (**Table 1**). All patients received either 400mg/day or 600mg/day of ribociclib for 5 days prior to a scheduled brain tumor resection. Six patients received five days of ribociclib (400mg once daily (QD)) plus everolimus (2.5mg QD) and underwent tumor resection at 2, 8 or 24 hours following the last dose. Six dose-escalation cohorts (n=3 each) reached a maximum dose-level of ribociclib (600mg QD) plus everolimus (70mg once weekly (QW), **Figure 1**). Both drugs were administered orally and well tolerated, and no dose-limiting toxicities were observed. The clinical outcomes for patients in this cohort are presented (**Extended Data Fig. 2b**), however, since no patient qualified for phase 1, clinical efficacy was not assessed.

**Figure 1.**
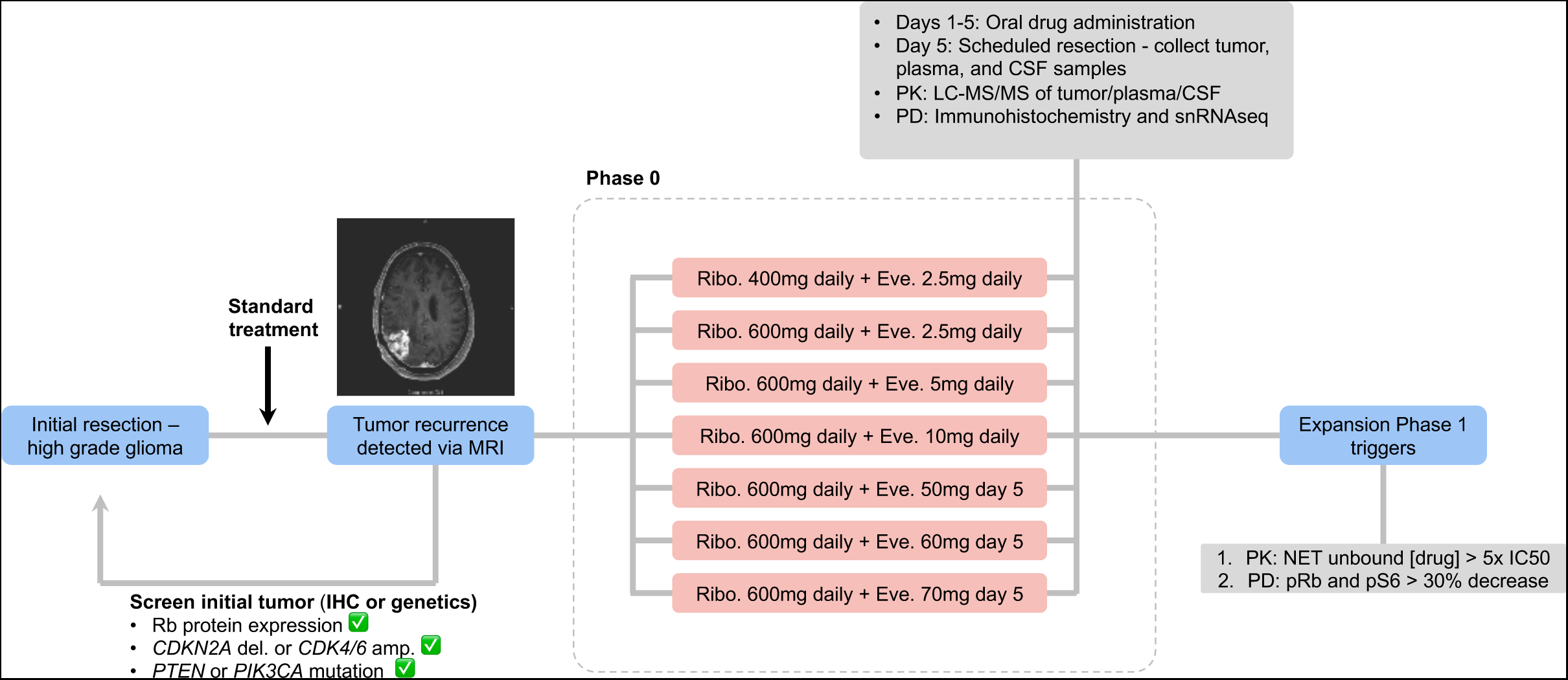
Phase 0/1 study design for ribociclib and everolimus in recurrent high-grade glioma. An overview of the clinical trial design and sample collection. Patients with a tumor recurrence detected via Magnetic Resonance Imaging (MRI) had their prior tumor screened for immunohistochemistry (IHC) and/or targeted genetic sequencing to select patients most likely to demonstrate a response to CDK4/6 and mTOR inhibition. As part of the phase 0 design, patients were allocated to 7 possible treatment arms including 400 or 600 mg ribociclib (Ribo.) daily for 5 days with dual inhibition of everolimus (Eve.) ranging from 2.5-10 mg daily for 5 days or a single higher dose on day 5 of treatment at 50 – 70 mg. Following the fifth day of drug exposure, the patients underwent a scheduled tumor resection. Tumor (Gadolinium contrast enhancing and non-enhancing regions), cerebrospinal fluid (CSF), and plasma specimens were assessed with a combination of pharmacokinetics (PK; tumor, plasma, and CSF) using liquid chromatography with tandem mass spectrometry and pharmacodynamics (PD; tumor) using IHC and single nucleus (sn) RNA sequencing. For patients to be eligible for the phase 1 expansion, the patient’s tumor tissue needed to meet pharmacokinetic/pharmacodynamic criteria for both ribociclib and everolimus. NET = non-enhancing tumor tissue.

**Table 1.** Clinical characteristics for ribociclib-everolimus phase 0 clinical trial cohort.

| **Characteristic** | **Phase 0 (n = 24)**<br>**recurrent tumors** |
| --- | --- |
| **Age at recurrent resection (years)** |  |
| Median (LQ-UQ) | 58 (46-65) |
| Unknown (n) | 0 |
| **Sex (%)** |  |
| Male | 17 (71%) |
| Female | 7 (29%) |
| **Tumor histology (%)** |  |
| Glioblastoma, IDH-wild-type | 21 (88%) |
| Astrocytoma, IDH-mutant, grade 4 | 1 (4%) |
| Astrocytoma, IDH-mutant, grade 3 | 2 (8%) |
| **IDH mutation status (%)** |  |
| IDH-wild-type | 21 (88%) |
| IDH-mutant | 3 (12%) |
| **Ribociclib dose (QD - 5 days)** |  |
| 400mg | 6 (25%) |
| 600mg | 18 (75%) |
| **Everolimus dose** |  |
| 2.5mg QD - 5 days | 9 (37.5%) |
| 5mg QD - 5 days | 3 (12.5%) |
| 10mg QD - 5 days | 3 (12.5%) |
| 50mg once on day 5 | 3 (12.5%) |
| 60mg once on day 5 | 3 (12.5%) |
| 70mg once on day 5 | 3 (12.5%) |

### Central nervous system pharmacokinetics and pharmacodynamics

To evaluate tumor pharmacokinetics and pharmacodynamics, samples were collected at the time of surgical resection, which followed 5 days of combined ribociclib and everolimus exposure. The following samples were collected for pharmacokinetic evaluation: tumor specimens from the Gadolinium enhancing tumor region, tumor specimens from non-enhancing tumor region, plasma, and cerebrospinal fluid (CSF). Total and unbound drug concentrations for ribociclib and everolimus in tumor, plasma, and CSF samples were determined using a validated liquid chromatography with tandem mass spectrometry (LC-MS/MS) method and equilibrium dialysis, as previously described [14]. Measuring unbound drug concentrations in the non-enhancing tumor regions is critical to assess drug penetration as non-enhancing tumor regions possess an intact blood brain barrier (BBB), are often unresectable, and a source of radiotherapy-induced neurological morbidity [15].

As previously seen in our monotherapy trial, there was robust CNS penetration of ribociclib as illustrated by total and unbound drug concentrations in tumor regions and CSF (**Extended Data Fig. 3a, Fig. 2a**) [8]. This was further supported by tumor- and CSF-to-plasma partition ratios for the total (Kp) and unbound drug (Kp, uu, **Extended Data Fig. 3b-c**). Following 5 days of exposure at the daily oral dose of 400 and 600 mg, ribociclib achieved a median unbound drug concentration of 170 nM (range, 65 – 1770 nM) and 634 nM (range, 68 – 2345 nM) in the non-enhancing tumor regions, respectively (**Supplementary Table 1**). These data were consistent with the unbound ribociclib concentrations in the non-enhancing tumor regions (median, 482 nM; range, 327 – 1295 nM) observed in ribociclib monotherapy study where the patients received a 5-day exposure at the daily dose of 900 mg [8]. Notably, we observed a significant dose-dependent increase in unbound ribociclib concentration in the contrast enhancing region (t-test, P = 0.02, **Figure 2a**) and a trend toward increased concentration in the non-enhancing regions and CSF (t-test, *P* = 0.18 and *P* = 0.06, **Figure 2a**) suggesting that the higher dose can deliver increased intratumoral concentration without dose-associated toxicities. The extent of ribociclib penetration in non-enhancing regions as assessed by Kp,uu was consistent across the different dose levels, with the median Kp,uu of 3.19, 3.20, and 2.00 at the daily dose of 400, 600, and 900 mg (monotherapy trial), respectively (**Supplementary Table 1, Extended Data Fig. 3c**). Pairwise comparisons of enhancing and non-enhancing regions from the same tumors highlighted the increased ribociclib concentration in enhancing regions and the importance of assessing non-enhancing unbound drug concentrations (**Extended Data Fig. 3d**, two-sided paired t-test, *P* = 0.03).

**Figure 2.**
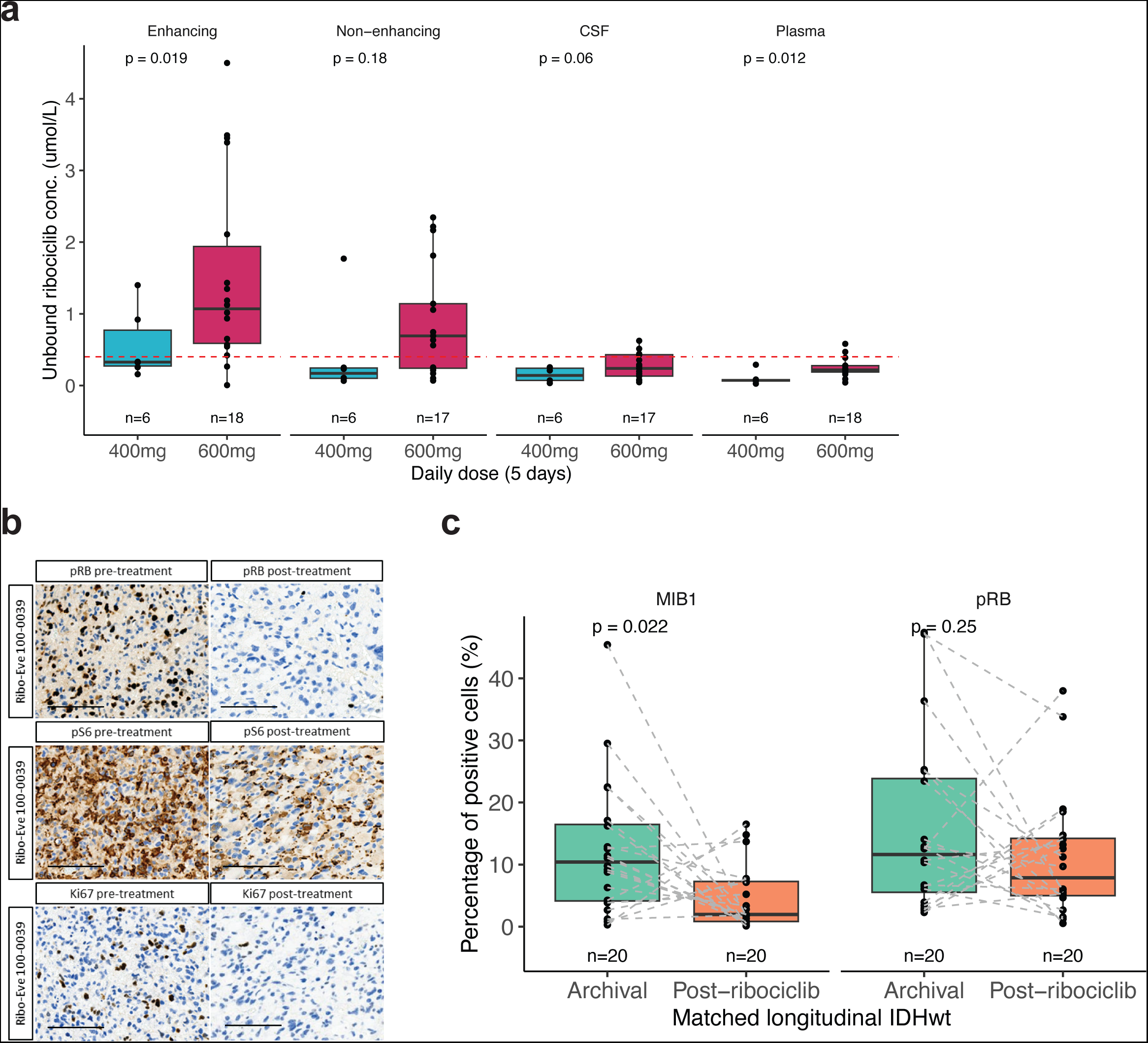
Pharmacokinetic and pharmacodynamic analyses of tumor samples exposed to ribociclib. **(a)** Unbound ribociclib concentrations in the Gadolinium enhancing tumor, non-enhancing tumor, cerebrospinal fluid (CSF), and plasma samples are presented for daily dose of 400 and 600 mg. Box plots in this figure and throughout the manuscript represent the median, lower and upper quartiles, and the whiskers indicates 1.5 times the interquartile range. Each point represents an individual measurement. Statistical differences were assessed with two-sided t-test. (**b**) Representative IHC images for pre-treatment (i.e., archival surgical sample) and post-treatment (phase 0 ribociclib exposure) for key pharmacodynamic markers in subject 100-0039. Scale bar, 100 microns. (**c**) Box plots represent the percentage of positive cells for each pharmacodynamic marker of ribociclib response including MIB1 (Ki67) and phosphorylated Rb. The dotted lines represent connections between samples collected from the same patient at the archival time point and following ribociclib phase 0 therapy. Statistical differences were assessed with a paired two-sided t-test.

CNS penetration of everolimus was evaluated at the daily oral dose of 2.5, 5, and 10 mg (given daily for 5 days), as well as at a single oral dose of 50, 60, and 70 mg (given on day 5). Overall, total everolimus concentrations and Kp indicated poor CNS penetration across different dose levels (**Extended Data Fig. 4a-b**, **Supplementary Table 2**). The median everolimus CSF concentration was 0.3 nM (range, below limit of quantification (BLQ) – 2.5 nM) across the different dose levels, indicating a negligible drug CNS penetration (**Extended Data Fig. 4a-b**, **Supplementary Table 2**). The unbound drug concentrations in both enhancing and non-enhancing tumor regions were below the lower limit of quantitation (< 0.1 nM), suggesting that inadequate levels of the pharmacologically active, unbound form of everolimus reach the tumor tissue in high-grade gliomas.

To assess the impact of these drugs on their targets, we next investigated the longitudinal stability of previously identified pharmacodynamic biomarkers of ribociclib response (MIB1, pRb, pFOXM1, and cleaved caspase 3) and activation of the mTOR pathway (p4EBP1, pS6) [8]. Given the subtype differences based on IDH status and small number of IDH-mutant samples (n = 3), we first restricted the longitudinal pharmacodynamic analysis to IDH-wild-type tumors. For IDH-wild-type patients with available pharmacodynamic data (n = 20), we observed a decrease in the proliferative MIB1 (Ki67) marker (paired t-test, *P* = 0.02) and a trend toward decreased phosphorylated Rb, which is a downstream target of CDK4/6 (paired t-test, *P* = 0.25, **Figure 2b-c, Supplementary Table 3**), for the surgical samples treated with ribociclib compared with their matched archival tissue samples. The reduction in MIB1 remained consistent when IDH-mutant tumors were included in the analysis (paired t-*test P* = 0.04, **Extended Data Fig. 5b**). There was a trend toward greater MIB1 reduction in tumors treated with higher ribociclib dose (median decrease 8.94% 600mg versus 2.93% 400mg, t-*test P* = 0.34, **Extended Data Fig. 5a**) suggesting that a higher ribociclib dose may produce a greater biological effect. Pathway activity of mTOR, as quantified by p4EBP1, pS6, was elevated but not significantly different at the surgical specimen compared to archival tissue suggesting variability in the upregulation of the mTOR pathway following CDK4/6 inhibition (paired t-test, *P* > 0.05, **Extended Data Fig. 5b**).

The requirements for expansion from the phase 0 treatment (5 days of treatment preceding surgical resection) to phase 1 were that both ribociclib and everolimus unbound drug concentrations needed to be five times greater than the biochemical IC_50_ (40nM for ribociclib, for 2 nM everolimus) and a greater than 30% decrease in both longitudinal pRb and pS6 levels (**Figure 1**). Accordingly, none of the patients in the phase 0 study qualified for phase 1.

### Single nucleus RNA sequencing of phase 0 tumor samples

To better understand the cellular changes that may occur in tumors due to active ribociclib exposure, we profiled 24 tumor samples with single nucleus RNA sequencing (10X Genomics, snRNAseq) including: 18 IDH-wild-type samples, 3 IDH-mutant, and 3 IDH-wild-type samples collected from a ribociclib monotherapy trial (NCT02933736) where 2 samples were longitudinally collected from the same patient during the phase 0 and phase 2 [8]. There were 87,888 cells remaining for analysis following application of quality control filters (**Extended Data Fig. 6a-b**). Unsupervised clustering followed by Uniform Manifold Approximation and Projection (UMAP) was used to identify the major tumor microenvironment cell types including neurons, oligodendrocytes, astrocytes, myeloid, lymphocytes, endothelial cells, mural cells, and malignant cells (**Figure 3a**). These cell type identities were confirmed by the high expression of key cell type-specific marker genes, including high expression of *CDK4*/*CDK6* in a subset of malignant cells (**Figure 3b**). We confirmed malignant cell status by inferring large-scale copy number alterations and identified 41,422 malignant cells (Methods, median 1,132 malignant cells per tumor, **Extended Data Fig. 6c-e**). We restricted subsequent analyses to tumors with a minimum of 25 malignant cells (**Extended Data Fig. 7a**). We next assigned malignant cell state by scoring each cell within tumor for the IDH-wild-type gene expression metaprograms previously identified in Neftel et al (IDH-wild-type only, **Figure 3c**) [10] assigning a malignant state based on highest relative metaprogram expression. We also determined whether a cell was actively in the cell cycle independent of the Neftel cell states by scoring for cell cycle metaprogram expression (Methods). To determine how snRNA metrics of cycling cells compared with immunohistochemistry based (IHC) proliferation markers, we correlated the proportion of malignant cells that were actively cycling in the snRNAseq data with the percent of MIB1 positive cells from matched IHC samples. We observed that these two metrics of proliferation were positively correlated with one another (Pearson’s correlation coefficient = 0.66, *p* = 0.005, **Figure 3d**) providing confidence in the snRNA cycling status assignment.

**Figure 3.**
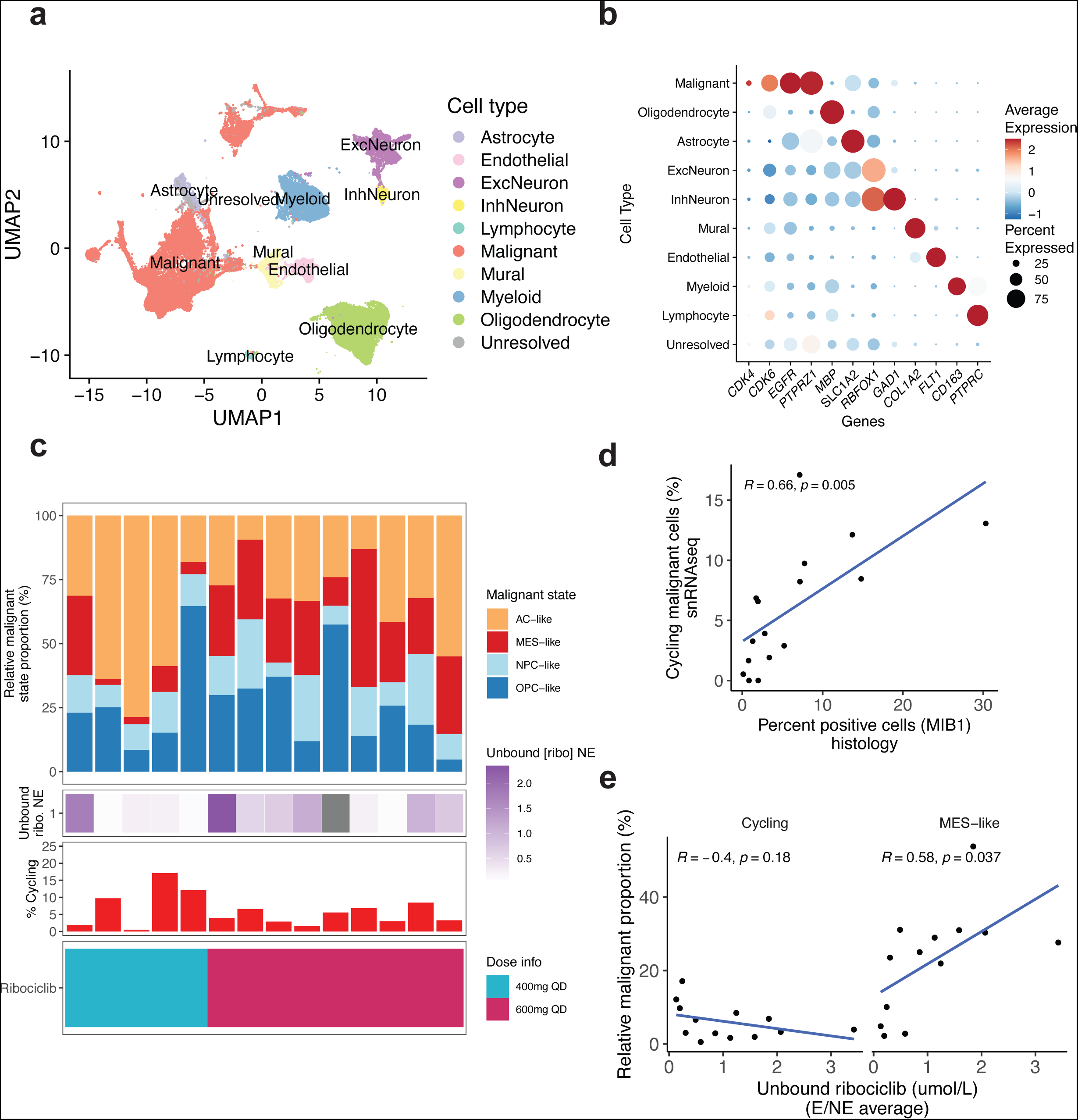
Single nucleus RNA sequencing (snRNAseq) of tumor samples treated with dual ribociclib and everolimus inhibition. (**a**) UMAP visualization for gene expression-based clusters that correspond to major cell types (n = 87,888 cells, n = 24 samples). Each point represents a single cell, and the colors denote cell type. Cell types were identified via integrated approach that considered de novo clustering identification, mapping to a non-malignant reference brain atlas, and inferred copy number alterations. (**b**) Dot plot represents the average normalized gene expression data per cell type for key marker genes. Dot color reflects expression scale and dot size reflects the percentage of cells annotated as a cell type expressing the displayed genes. Genes shown represent selected genes from Seurat’s FindMarkers for each cell type as well as *CDK4*/*CDK6* expression. (**c**) Stacked bar plots represent the relative malignant proportions assigned to each Neftel cell state in the top panel (n = 14 IDH-wildtype samples from phase 0 ribociclib-everolimus cohort). The middle top panel reflects the unbound ribociclib concentration measured in the contrast non-enhancing region of the tumor. The middle bottom panel represents the proportion of malignant cells that are actively cycling according to snRNA. The bottom panel represents the daily ribociclib oral dose (5-days preceding resection). (**d**) For samples with sufficient snRNA cells passing quality control and IHC, the Pearson correlation between histological estimates of cycling cells and cycling malignant cells in the snRNA data is presented (n = 16, IDH-wildtype + IDH-mutant). (**e**) The Pearson correlation coefficient is presented for the average (non-enhancing and enhancing) unbound ribociclib concentration (umol/L) and the relative proportion of MES-like malignant cells (n = 13 IDH-wildtype from ribociclib-everolimus cohort).

We then investigated whether active ribociclib exposure may be linked with differences in malignant or microenvironmental cell states by testing the association for the proportion of each malignant cell state with the unbound ribociclib concentration in the enhancing and non-enhancing regions (phase 0 trial samples only). We examined the average ribociclib concentration across the contrast enhancing or non-enhancing regions since we did not use neuronavigation techniques to collect tissue for snRNA profiling (Methods). The correlation between increased unbound ribociclib and malignant states revealed a positive association with increased MES-like proportions (Pearson’s correlation coefficient = 0.58, *p* = 0.04) and a trend toward decreased cycling proportions (Pearson’s correlation coefficient = −0.4, *p* = 0.18, **Figure 3e, Extended Data Fig. 7b**). Increased MES-like abundance has previously been associated with the tumor immune microenvironment [10] and may also represent an adaptation to therapeutic challenges [9, 16]. To better understand the observed association between increased MES-like with ribociclib concentration, we tested whether ribociclib tumor concentration was correlated with the relative proportion of tumor microenvironment cell compartments. There was a positive correlation between unbound ribociclib and immune compartment abundance (myeloid + lymphocytes) (Pearson’s correlation coefficient = 0.49, *p* = 0.04, **Extended Data Fig. 7c**) suggesting that microenvironments with greater immune cells may be more permissive to drug penetration. Together, these data suggest that increasing pharmacologically active ribociclib levels are associated with shifts in malignant and microenvironmental cell states.

### Malignant cell state shifts are associated with ribociclib exposure

To expand our understanding of ribociclib’s impact on intratumoral malignant cell state diversity, we next sought to compare the relative malignant cell states with an independent cohort of recurrent IDH-wild-type tumors that received the standard of care therapy (i.e., the alkylating agent temozolomide, radiation, and surgery) but were not exposed to neoadjuvant ribociclib prior to the recurrent tumor resection [9]. **Supplementary Table 4** shows a comparison of the standard treatment and phase 0 cohorts based on subject demographics. Overall, 31 recurrent GBM that received standard therapy without neoadjuvant ribociclib exposure were analyzed for snRNAseq (Methods).

We first assessed whether there were any shifts in malignant cellular state distributions between the standard treatment cohort and ribociclib following standard treatment cohort. We observed that a lower proportion of malignant cells that were actively cycling (two-sided t-test, *p* = 2.0E-04) in those exposed to ribociclib providing orthogonal support to the observation that ribociclib suppresses cell proliferation *in vivo* (**Figure 4a**). We also found a decrease in the neural progenitor-like (NPC-like) state for the ribociclib-exposed samples (two-sided t-test, p = 0.002, **Figure 4a**), which remained significant when restricting to non-cycling cells (**Figure 4b**). The NPC-like state has been previously associated with *CDK4* amplifications, and our results suggest that inhibitors of CDK4 selectively downregulates the NPC-like expression program [10]. Among the ribociclib monotherapy samples, we longitudinally profiled a patient’s tumor from both a phase 0 (5-day drug exposure) and phase 2 (104 days) using snRNAseq. While only a single case, we did observe a reduced fraction of NPC-like malignant cells in the phase 2 sample (**Extended Data Fig. 8a**).

**Figure 4.**
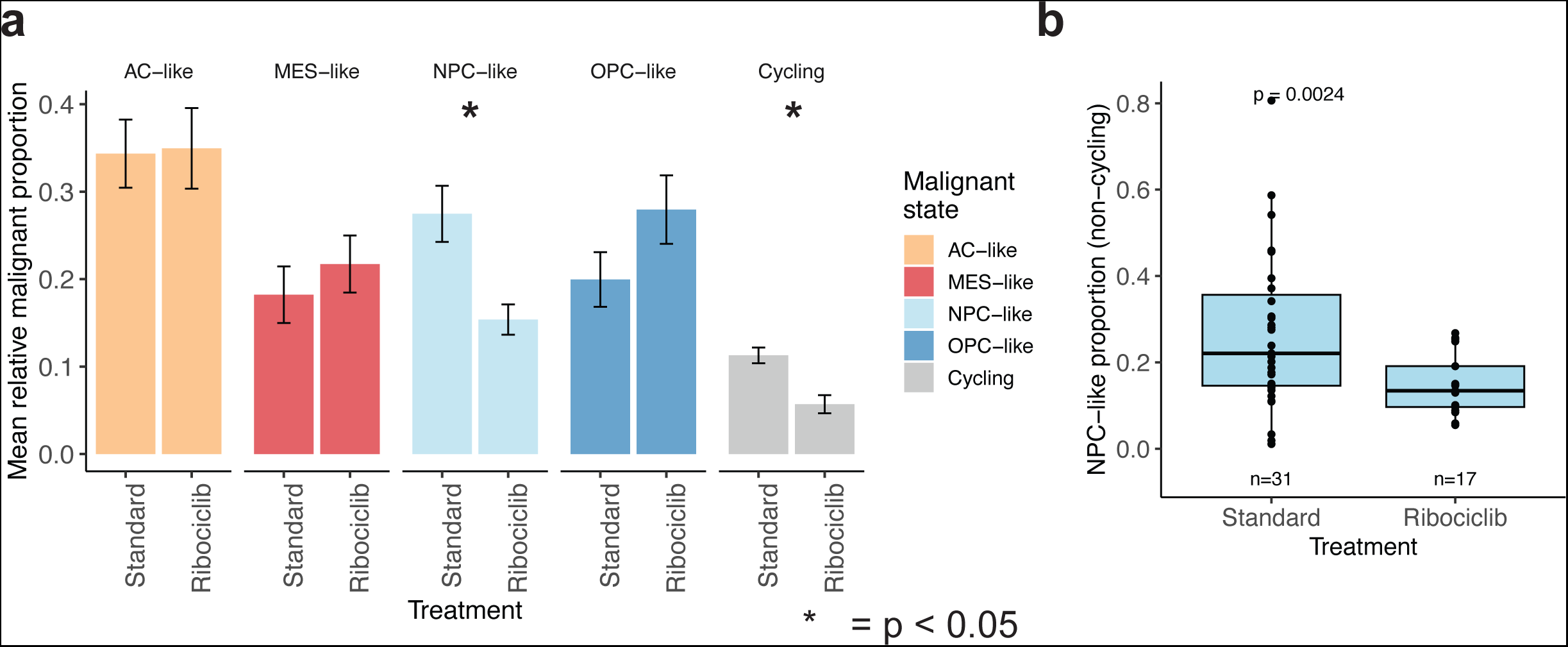
Malignant cell state distribution differences in standard treatment versus neoadjuvant ribociclib following standard treatment cohorts. (**a**) The mean relative malignant cell proportion is presented for the samples treated with standard of care therapy (n = 31 snRNA IDH-wild-type recurrences) and samples treated with neoadjuvant ribociclib that followed previous standard therapies (n = 17 IDH-wild-type recurrences). The four malignant cell states (AC-like, MES-like, NPC-like, OPC-like) plus the proportion of malignant cells that are actively cycling are shown. The bar height represents the mean value per group and the bars represent +/- the standard error. Statistical difference was assessed with two-sided t-test. Asterisk (*) indicates comparison below the threshold for statistical significance (*P* < 0.05). (**b**) Box plot with individual points representing observations for each tumor sample for the NPC-like population that were not actively cycling. That is, the relative malignant proportion for only non-cycling malignant cells.

We next assessed whether there were any differences in the tumor microenvironment (TME) abundance across cohorts since the TME plays a role in shaping malignant cell state abundance [10]. We observed significantly greater oligodendrocyte abundance in the standard treatment versus ribociclib-exposed cohort and no significant differences in the other major cell types (**Extended Data Fig. 8b**). Importantly, we confirmed that NPC-like malignant state abundance was not associated with TME cell type abundance suggesting that the observed cohort difference in NPC-like abundance was not likely driven by cohort differences in the local microenvironment (**Extended Data Fig. 8c**). Together, these comparative analyses across cohorts support the anti-proliferative effect of ribociclib in IDH-wild-type gliomas and suggests that ribociclib exposure may be associated with a smaller NPC-like malignant state compartment.

### *In vitro* ribociclib exposure depletes cycling and NPC-like cell states

To confirm whether ribociclib exposure selectively depletes or enriches for a particular malignant state, we exposed patient-derived recurrent glioblastoma cell lines to ribociclib or vehicle control (DMSO) *in vitro* (**Figure 5a**). These patient-derived cell lines (GB126, GB239, and GB86) were selected based on similar genomic profiles to the eligibility criteria (e.g., all had a high-level *CDK4* amplification, **Extended Data Fig. 9a**) and included one cell line (GB239) that was derived from resected tumor tissue in the phase 0 trial (100-0039). To establish the selected ribociclib dose for each cell line, we exposed cells to ribociclib for 5 days across a dose range and selected the dose per cell line that achieved a 50% inhibition of pRb (Methods, **Extended Data Fig. 9b**, dose 0.5-5uM). Interestingly, the sample with the highest *CDK4* copy number amplification level that did not harbor other focal oncogene amplifications (GB126) was most sensitive to CDK4/6 inhibition. We next performed single cell RNAseq on the ribociclib-treated (n = 3 per cell line) and DMSO control samples (n = 3 per cell line). Following filtering for quality cells (138,870 cells post-quality control, n = 18 samples), we performed dimensionality reduction and clustering (**Extended Data Fig. 9c**). This revealed cell line dependent and treatment-enriched clusters suggesting a treatment effect (**Extended Data Fig. 9d**). We found that *CDK4* gene expression was reduced but remained high following ribociclib treatment, consistent with ribociclib binding to CDK4 to inhibit its function (**Extended Data Fig. 9e**).

**Figure 5.**
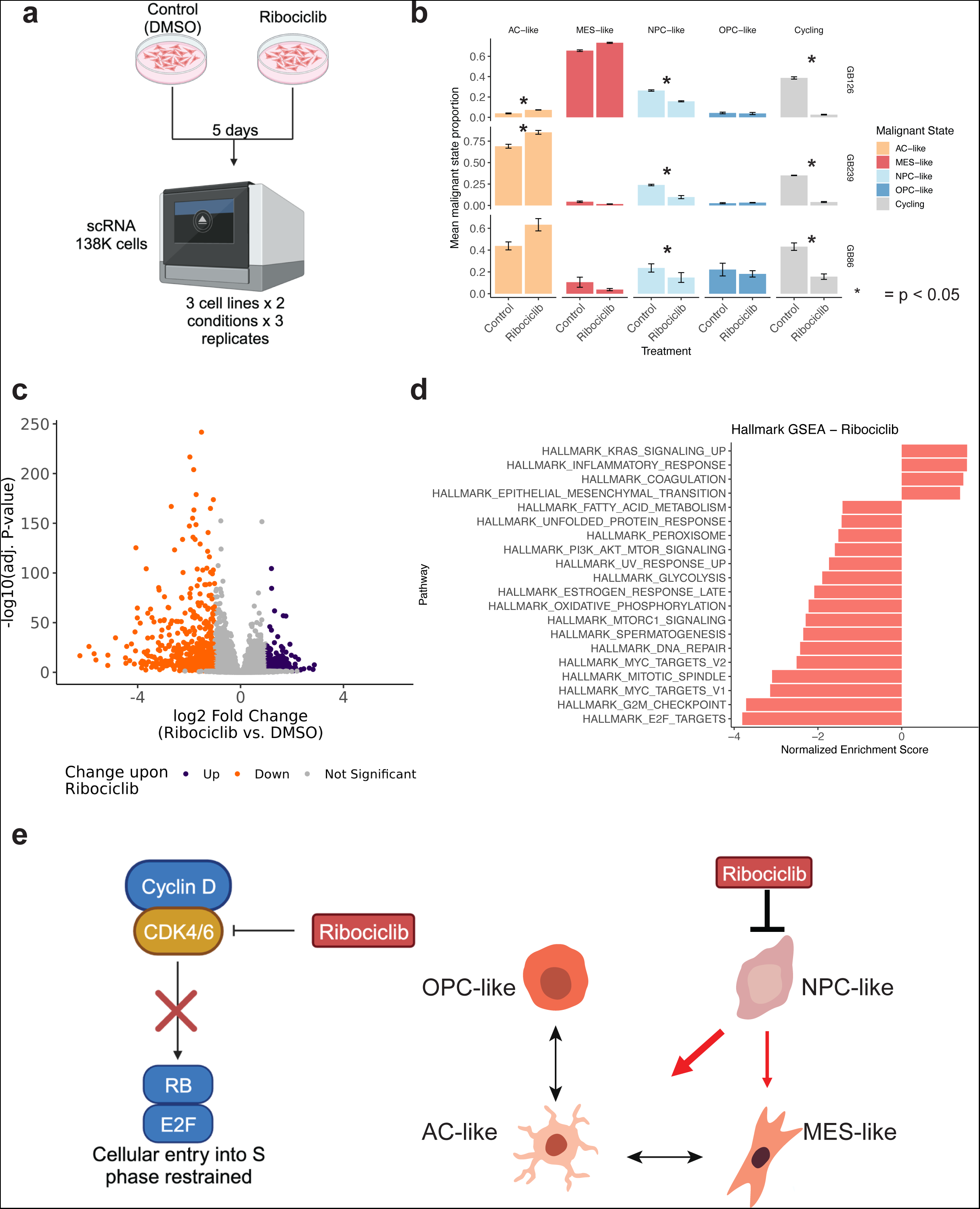
*In vitro* pharmacological perturbation of malignant cell states. (**a**) Experimental design for ribociclib *in vitro* perturbation experiments. Patient-derived cells were split and allocated to either control (DMSO) or ribociclib treatment for 5 days at empirically defined doses that inhibit pRb to 50% per cell line. Cells were then profiled by single cell RNA sequencing. Each experiment was performed with 3 biological replicates. (**b**) The relative proportions of each malignant state are shown per cell line. The height indicates the mean value and the bars +/- the standard error. Statistical assessment was made with a paired two-sided t-test. Asterisk (*) indicates comparisons below the P < 0.05 threshold. (**c**) Volcano plot represents the pseudobulk differential gene expression analysis. Each point reflects an individual gene with point color reflecting upregulation following ribociclib exposure (purple) and downregulation (orange) based on absolute(log2 FC) > 1 and adjusted p-value < 0.05. Y-axis indicates the −log10(adjusted p-value). (**d**) Gene set enrichment analysis (GSEA) for differentially expressed genes between ribociclib exposure and DMSO. Bars show normalized enrichment score where values less than 0 would reflect underrepresentation in the differential expression rank (e.g., E2F targets downregulated in ribociclib exposure). (**e**) Ribociclib’s impact on cellular states in high-grade glioma. Ribociclib penetrates into glioma tissues to inhibit CDK4/6 activity leading to hypo-phosphorylation of Rb and restrained entry into the cell cycle (left panel). Ribociclib exposure shifts malignant states away from NPC-like populations to more differentiated cell states (e.g., AC-like and MES-like, right panel).

We then scored the cells for malignant state gene expression metaprograms (Neftel states) and examined their relative proportions across treatment groups in cell line passage-dependent manner (Methods, **Figure 5b**). Across all three cell lines, we found a consistent decrease in the cycling cells (paired t-test, *P* = 8.7E-4 GB126, *P* = 5.4E-4 GB239, *P* = 0.04 GB86) consistent with cytostatic effect due to cell cycle arrest [17]. The relative NPC-like malignant state proportion was also reduced for all three cell lines (paired t-test, *P* = 4.3E-4 GB126, *P* = 0.02 GB239, *P* = 0.038 GB86) with a compensatory increase towards AC-like (paired t-test, *P* = 6.6E-3 GB126, *P* = 0.03 GB239, *P* = 0.06 GB86) and in one cell line the MES-like state (GB126), both of which represent more differentiated-like malignant cells (**Figure 5b**) [12]. These cell state changes were not accompanied by changes in broad inferred chromosomal gains or losses events (**Extended Data Fig. 10a**). The cell line-specific effects toward greater AC-like and, in one case, also greater MES-like may reflect genetic differences in these cell lines (**Extended Data Fig. 9a and 10a**) that increase the likelihood of transition toward a specific differentiated-like state (e.g., *EGFR* amplification associated with increased AC-like in GB239). Additionally, when we restricted our analyses to those cells not actively cycling, we observed a consistent decrease in NPC-like (paired t-test, *P* = 1.4E-06), increase in AC-like (paired t-test, *P* = 2.6E-03) supporting this pharmacological driven cell state shift can occur independent of cell cycle (**Extended Data Fig. 10b**).

To identify broader patterns of ribociclib activity, we performed pseudobulk differential gene expression analyses where cells from a sample are collapsed into a single gene expression profile to perform cell state independent analyses. This analysis tested the effect of treatment while adjusting for experimental replicate and identified 678 upregulated and 582 downregulated genes (adj. p < 0.05, abs(log2FC) > 1, **Supplemental Table 5**) in the ribociclib-treated samples (**Figure 5c**). A gene set enrichment analysis on significantly downregulated genes in the ribociclib treated cells identified cell cycle as the most affected hallmark pathway (**Figure 5d**). In contrast, the upregulated genes were enriched for programs including inflammatory response and epithelial-to-mesenchymal transition nominating candidate programs that may drive resistance to ribociclib. Together with our phase 0 *in vivo* results, these *in vitro* findings highlight ribociclib’s ability to modify glioma cellular state distribution via cell cycle downregulation and a shift away from the neural progenitor state to more differentiated malignant cell states (**Figure 5e**).

## DISCUSSION

In this dual-drug phase 0 trial with a pharmacokinetic/pharmacodynamic-triggered phase 1 expansion, we provide a template for an innovative trial design that allows dose-escalation and rapid, early-stage evaluation of pharmacokinetics, pharmacodynamics, and molecular impacts of two targeted agents, ribociclib and everolimus. Importantly, this design incorporated single cell-level transcriptional analyses to comprehensively characterize tumor samples on active therapy. Beyond confirmation of effective ribociclib penetration and ineffective everolimus penetration into both contrast enhancing and non-enhancing tumor regions, our study provides critical insights into ribociclib’s ability to exert significant anti-proliferative effects on glioma cells, as evidenced by reduced cell proliferation markers and shifts in malignant cell states towards more differentiated phenotypes. Our study also suggests that a higher ribociclib dose may achieve a greater biological effect without any observable toxicity. In contrast, everolimus’ lack of CNS penetration provides support that its use in adult glioblastoma therapy should not be further pursued. Together, our combined *in vivo* and *in vitro* results provide proof-of-concept that a drug that therapeutically targets malignant cell states can penetrate glioma tumor tissue and have a biological effect.

The integration of snRNA-seq technology into the phase 0 study design provided an unbiased insight into the impact of targeted therapies on different cellular states within the tumor milieu. While pharmacodynamic analysis of changes in expression of proximal markers (e.g., pRb or pS6) provides information on target inhibition to some extent, snRNA-seq analysis provides broader information on overall cell state and cell cycle regulation. For example, our analyses identified molecular effects of ribociclib on cell cycle regulation as evidenced by decreased proliferating malignant and neural progenitor cell-like (NPC-like) populations, among which the latter could not have been detected by traditional pharmacodynamic analyses [10]. Our observations of smaller cycling and NPC-like populations were confirmed and extended in our *in vitro* perturbation analyses where the shift away from the NPC-like cell state was coupled with a concomitant increase in more differentiated malignant states (AC-like and MES-like). The ability to pharmacologically induce a differentiation effect in glioma cells was recently shown in IDH-mutant tumors that were treated as part of a phase 1 clinical trial with a mutant IDH inhibitor, examined with snRNAseq, and compared with publicly available IDH-mutant single cell reference profiles [18]. Together, the mutant IDH inhibitor trial and our CDK4/6 inhibitor trial’s results highlight the utility of using single cell genomics to profile glioma tissue samples on trial to understand how drugs function *in vivo*. Future studies using these approaches may help identify patients who are most likely to respond to targeted inhibitors and to combination therapies. More broadly, these studies also suggest that pharmacologically shifting malignant cells towards a more differentiated malignant state may restrict glioma cell plasticity by depleting progenitor populations and slow tumor growth via cell cycle inhibition [19]. Nevertheless, despite the reduction in proliferating and the NPC-like stem/progenitor populations, we have previously observed that ribociclib monotherapy, in a small cohort, had limited clinical benefit at recurrence compared with historical cohorts [8]. Hence, future studies will need to examine whether targeted inhibition can be more effective when CDK4/6 inhibitors are given earlier in a treatment schedule and whether predictors of response can be identified. For example, recent preclinical work identified that recurrent IDH-mutant glioma models are sensitized to CDK4/6 inhibitors by *CDKN2A*/*B* deletion [20]. Future clinical trial designs that incorporate genomic profiling and non-invasive longitudinal sampling (e.g., cerebrospinal fluid) may provide both more precise patient selection and an ability to continuously monitor treatment response.

While ribociclib showed robust penetration and a biological effect, our study found that no unbound everolimus was detected in the glioma tissue across six dose escalation levels. These data suggest no future clinical potential for everolimus in adult glioblastoma therapy and raise important pharmacokinetic and pharmacodynamic questions regarding its known therapeutic effects in subependymal giant cell astrocytoma (SEGA) in patients with tuberous sclerosis [21]. Given the importance of mTOR signaling pathway in glioma pathogenesis, alternate strategies are needed to modulate the mTOR pathway more effectively within the glioma microenvironment. This might include development of novel brain penetrant mTOR inhibitors or drug delivery strategies that can circumvent BBB. An ongoing phase 0 trial (NCT05773326) is evaluating the pharmacokinetic/pharmacodynamic effects of superselective intra-arterial infusion as a means of bypassing the BBB to deliver the mTOR inhibitor, temsirolimus, in recurrent high-grade glioma patients. If successful, this delivery technique could significantly improve the bioavailability of therapeutic agents like everolimus within the glioma microenvironment.

The study design used in this trial allowed investigation of two targeted therapies concomitantly with dose de-escalation and escalation rules to accommodate challenges with drug toxicities and unsuitability of microdosing due to BBB. However, there were some study design limitations. Unlike conventional phase 0 studies in the medical oncology field, the comparator for pharmacodynamic response in phase 0-collected tissue were archival tissues from a previous surgery and not pre-treatment biopsies. Collecting adequate pre-treatment glioma tumor tissue required for molecular analyses such as IHC and snRNA remains a challenge in evaluating glioma patient treatment response where additional biopsies may pose an increased risk to the patient [3, 22]. Lack of pre-treatment biopsy tissue affected our ability to utilize the most appropriate comparator for the phase 0-treated tissues. Furthermore, the absence of fresh frozen archival tissue matched to phase 0 tissues prevented us from performing longitudinal single cell analyses on the same tumor samples where it is not yet possible to perform this analysis in formalin fixed tissues. To address this limitation, our single cell analysis of tumor samples that had previously received standard of care and were actively on ribociclib were compared with single cell data obtained from recurrent glioma samples following standard of care treatment. Ongoing Phase 0 studies are making an effort to incorporate pre-treatment stereotactic needle biopsied tissue as a means of improving comparator tissue.

In summary, our phase 0/1 trial of ribociclib and everolimus offers valuable insights into the therapeutic potential of CDK4/6 inhibition for high-grade gliomas. The insights gained from single cell analyses have provided a deeper understanding of ribociclib’s impact on malignant cell states, offering a promising avenue for future therapeutic interventions. While ribociclib shows promise in modulating malignant cell states, the challenges with everolimus highlight a critical need for advanced strategies in drug delivery and mTOR pathway targeting. Continued investigation into regulators of malignant cell state and transitions will be crucial in designing effective combination therapies for glioma. As the neuro-oncology field moves forward, the integration of innovative trial designs, molecular profiling techniques, and non-invasive longitudinal sampling methods will be pivotal in accelerating the development of therapies for glioma that improve patient outcomes.

## Data Availability

Gene expression count matrices processed by cellranger will be made available via the Gene Expression Omnibus (accession number available once deposited) and analysis code will be made available via github upon publication.

## Acknowledgements

We thank the patients who enrolled and participated in this study. This work was funded by the Ben and Catherine Ivy Foundation and the Barrow Neurological Foundation. We gratefully acknowledge the contribution of the Single Cell Biology service, the Genome Technologies service, and Cyberinfrastructure high performance computing resources at The Jackson Laboratory for expert assistance with the work described herein. These shared services are supported in part by the JAX Cancer Center (P30 CA034196). This work was supported by NIH grants R01 CA237208, R01 CA271601, OT2 CA278649 and U24 CA264379 (R.V). Schematic images were created with BioRender.com.

## Author contributions

Project conception and design: A-C.T., S.M., R.V., N.S.

Acquisition of data (patient engagement, pharmacokinetic/pharmacodynamic, snRNA): A-C.T., J.J., J.M., Y-W.C., C.M., A.D., L.E., Y.F., S.K., A.H., J.H., A.T., J.L.

Tissue handling and cell extractions (sc/snRNA): P.G., E.C., J.M.

Analysis and interpretation of data: K.C.J., A.S., W.F., A.H., A-C.T., J.L., S.M

Writing, review, and revision of manuscript: K.C.J., A-C.T., S.M., R.V., N.S.

Study supervision: A-C.T., S.M., R.V., N.S.

## METHODS

### Sample acquisition and study design

This phase 0/1 clinical trial (NCT03834740) was conducted at the Ivy Brain Tumor Center at the Barrow Neurological Institute/St. Joseph’s Hospital and Medical Center in Phoenix, AZ. Patient enrollment spanned from 2019-2022. The study had the following inclusion criteria: 1. Patients needed to be 18 years or older; 2. Prior resection of a histologically diagnosed grade III or IV glioma (2016 World Health Organization diagnostic criteria) that progressed following standard therapy (i.e., Stupp regimen of maximally safe surgical resection, temozolomide, and fractionated radiotherapy [1]); 3. The tumor recurrence needed to be confirmed with Gadolinium contrast-enhanced magnetic resonance imaging (MRI) or diagnostic biopsy followed by pathologic review; 4. Archival tissue needed to demonstrate one of the following: a) Rb protein positivity on immunohistochemistry (>= 20%) or the absence of *RB1* mutations; b) chromosomal loss of *CDKN2A*/*B*/*C* or *CDK4*/*6* or *CCND1*/*2* amplification on microarray-based comparative genomic hybridization; c) mTOR positive with *PTEN* loss or *AKT3* amplification or mutations for *PIK3CA* or *PIK3R1*, or pS6 positivity (>=10%); 5. Patients needed to voluntarily agree to participate by signing a written informed consent document; 6. Patients needed to be able to swallow ribociclib and everolimus capsules/tablets. The study received approval from the institutional review board at St. Joseph’s Hospital and Medical Center/Dignity Health and was conducted in accordance with the Declaration of Helsinki and Good Clinical Practice.

Enrolled phase 0 patients were administered 400-600 mg QD of ribociclib and 2 mg QD to 70mg QW of everolimus for 5 days preceding a planned cranial tumor resection (**Figure 1**). Patients were assigned to dose escalation arms that included:

1. 400 mg ribociclib QD + 2.5 mg everolimus QD (n = 6)
2. 600 mg ribociclib QD + 2.5 mg everolimus QD (n = 3)
3. 600 mg ribociclib QD + 5 mg everolimus QD (n = 3)
4. 600 mg ribociclib QD + 10 mg everolimus QD (n = 3)
5. 600 mg ribociclib QD + 50 mg everolimus QW (n = 3)
6. 600 mg ribociclib QD + 60 mg everolimus QW (n = 3)
7. 600 mg ribociclib QD + 70 mg everolimus QW (n = 3)

Pre-operative MRI and intra-operative neuronavigation were used to collect tumor specimens from both the contrast-enhancing and non-enhancing regions. Blood plasma and cerebrospinal fluid (CSF) were also collected from patients at time of surgical resections. These specimens were used for pharmacokinetic and pharmacodynamic analyses. Resected tumor tissue for 21 specimens was immediately frozen to process for single nucleus RNA sequencing. In two cases, patients that received ribociclib-everolimus therapy (5 days) were later deemed to have presented with treatment-associated pseudo-progression following surgical resection. These patients were replaced on the trial for pharmacokinetic and pharmacodynamic analyses; however, these tumor samples were profiled with snRNA following the drug treatment and resection. One pseudo-progression snRNA sample was excluded due to low malignant cell count, while the other was retained due to malignant cell detection. The resected tissue collected for snRNA analyses did not have annotation for contrast enhancing versus non-enhancing regions.

### Ribociclib and everolimus pharmacokinetics

Pharmacokinetic evaluation was conducted as previously described [8] using a validated liquid chromatography with tandem mass spectrometry method [14]. Briefly, blood samples were collected, and plasma separated from whole blood via centrifugation (4 C 1500 x g for 10 minutes). Tumor samples collected from contrast-enhancing and non-enhancing regions were rinsed with cold PBS to remove any residual blood, dried, and subsequently snap frozen. CSF samples were intraoperatively collected.

### Ribociclib and everolimus pharmacodynamics

Pharmacodynamic biomarkers for ribociclib were assessed using immunohistochemistry (IHC) with the following antibodies: anti-pRb (Cell Signaling Technology; #8516, 1:400), anti-pFOXM1 (Cell Signaling Technology; #14655, 1:200), anti-MIBI1 (DAKO; M724029, 1:100), and anti-cleaved caspase 3 (Cell Signaling Technology; #9661, 1:300). The pharmacodynamic biomarkers for everolimus were assessed using IHC with the following antibodies: anti-p4EBP1 (Cell Signaling Technology; #2855, 1:500) and anti-pS6 ribosomal protein (Cell Signaling Technology; #4858, 1:100). Stainings were performed using standard immunohistochemistry protocols with the BOND RX automated system (Leica Biosystems, Wetzlar, Germany). The stained slides were imaged using the Leica Versa microscope and analyzed using Aperio Image analysis software. Percentage of positive cells were quantified from random selection of at least 12 regions of interest for each tissue section.

### Single nucleus RNA sequencing tumor samples

Available frozen clinical glioma specimens were obtained from patients that had received combination therapy of ribociclib and everolimus (*n* = 21 samples) or ribociclib monotherapy (n = 3 samples). Frozen glioma specimens (n = 3) from a ribociclib monotherapy trial at St. Joseph’s Hospital and Medical Center in Phoenix, AZ (NCT02933736) were analyzed to expand the dataset of samples treated with ribociclib [8]. Nuclei isolation for all samples was performed within the same laboratory using an assay optimized for the isolation of nuclei from non-diseased brain and brain tumor tissues (protocols.io: dx.doi.org/10.17504/protocols.io.81wgb657ylpk/v1). Briefly, tissue samples were thawed and mechanically dissociated in Nuclei EZ lysis buffer (Millipore Sigma) via dounce homogenization. The solutions were incubated on ice for 5 minutes and mixed 1-2 times during incubation. Single-nuclei suspensions were filtered through a 70 μm strainer and centrifuged at 500g for 5 minutes at 4 C, resuspended in Nuclei EZ Lysis Buffer and incubated on ice for 5 minutes. The solutions were centrifuged at 500g for 5 minutes at 4 C and resuspended in 1% BSA/0.2 UμL RNase Inhibitor/PBS buffer (three times). For the final resuspension, the DAPI was added to the buffer, the solution was filtered through a 40 μm strainer, cells were counted on a Countess II automated cell counter (Thermo Fisher Scientific), and nuclei were taken into the 10x Genomics (3’ version 3 chemistry) workflow with a targeted recovery of 6,000 nuclei.

### Single nucleus RNA sequencing analysis

Cellranger count (6.1.2) function was used to align sequencing reads from FASTQ files to the GRCh38-2020-A reference transcriptome with the parameter include introns set to true. Count matrices were then loaded into R using Seurat’s (version 4.3.0) Read10X function. Cells were filtered out based on the following quality control criteria: the number of genes detected needed to be greater than 500 and fewer than 10,000 with a maximum percentage of mitochondrial genes of 5%. The DoubletFinder software was used to identify potential nuclei doublets (i.e., instances where multiple nuclei occupy the same droplet), which were then filtered out [23]. The expression data was then processed by normalization, scaling, PCA based on genes differentially expressed across brain cell types, Harmony batch correction for sample preparation batch [24], Uniform Manifold Approximation and Projection (UMAP), and clustering using the Louvain algorithm with resolution set to 0.6. Cell type annotation was performed based on gene expression markers for each cluster and confirmed via mapping to a normal reference brain atlas (Azimuth) and the software inferCNV of the Trinity CTAT Project (https://github.com/broadinstitute/inferCNV) was used to confirm malignant status based on the presence of copy number alterations. Briefly, the reference cells were set to myeloid and oligodendrocytes from that tumor sample and in cases where these cells were not detected in sufficient number, endothelial cells. Predicted copy number altered regions based on a moving average of a 101 gene window were determined with the Hidden Markov Model (HMM) parameter enabled. The HMM predicted copy number alterations levels were extracted to define malignant and non-malignant cells based on the following approach. Cells assigned to a cluster consistent with malignant marker expression that also possessed greater than 25% of chromosome 7 inferred gain and/or 25% of chromosome 10 inferred loss (two hallmark alterations in high grade gliomas [4, 5, 25]) were retained as malignant. If a putative malignant cell did not meet this threshold, it was labelled as unresolved and excluded from downstream analyses. Similarly, if a cell assigned to a non-malignant gene expression cluster had 25% of chromosome 7 inferred gain and/or 25% of chromosome 10 inferred loss, then it was labelled as unresolved and excluded from downstream analyses. Samples that did not have a minimum of 25 malignant cells were excluded from analysis of malignant states. Seurat’s AddModuleScore function was used to score malignant cells separately within each tumor sample for the relative expression of each Neftel cell state (i.e., AC-like, MES-like, OPC-like, NPC-like) and cell cycle against a background set of genes [10]. Cell state assignment was then determined based on the Neftel cell state with the highest relative expression. Cell cycle status was assigned in independent of the Neftel cell state such that each cell would be classified as AC-like, MES-like, OPC-like, or NPC-like along with its cell cycle status – cycling or not.

### Publicly available single nucleus RNA sequencing data cohort

Raw gene expression counts from single nucleus RNA sequencing (10x Genomics) for longitudinally collected primary and recurrent glioblastoma samples were retrieved from GSE174554. Notably, only recurrent samples were analyzed for differences with ribociclib-everolimus treated cohort. All subjects included in this cohort were noted to have received standard of care (chemotherapy and radiotherapy) and were restricted to IDH-wild-type tumors [9]. We reprocessed the raw gene expression count data for IDH-wild-type gliomas and analyzed the cellular states in the manner described for the phase 0 ribociclib-everolimus samples.

### Determination of ribociclib dose *in vitro*

Patient-derived glioma stem cell lines (GSCs; GB126, GB86, and GB239) were established from resected GBM tumor tissue at the St. Joseph’s Hospital and Medical Center/Barrow Neurological Institute. Cell lines were genetically profiled for mutations and copy number aberrations using the IvySeq custom gene panel established at the pharmacodynamics core at the Ivy Brain Tumor Center. All cell lines harbored high-level *CDK4* amplifications among other alterations. GB239 was derived from a tumor sample included in this phase 0 trial (tumor sample: 100-0039). All human GSCs were cultured as described previously [11]. GSCs were cultured as spheres on non-tissue culture-treated 10cm plates or as adherent cultures on laminin on tissue culture-treated 10 cm plates (ThermoFisher Scientific). GSCs were grown in DMEM/F12 media, supplemented with B27, N2 (Invitrogen, ThermoFisher Scientific) 1% penicillin-streptomycin in the presence of 20 ng/ml epidermal growth factor (EGF) and basic fibroblast growth factor (bFGF) (MilliporeSigma). GB126, GB86, and GB239 were treated with 0.5μM, 1μM, and 5μM concentration of ribociciblib and incubated for five days to determine ribociclib dose per cell line. Cellular protein from cultured cells were homogenized in RIPA lysis buffer containing protease and phosphatase inhibitors (ThermoFisher Scientific), rotated at 4 °C for 20 minutes and then centrifuged at 15,000 rpm for 10 minutes at 4 °C. Protein concentration from whole-cell extracts were determined using the Bradford Protein Assay (ThermoFisher Scientific). Equal amounts of protein (10-40 μg/lane) were loaded onto a 10% or 12.5% SDS-PAGE gels and transferred to a polyvinylidene fluoride membrane (PVDF; Millipore-Sigma). Membranes were blocked with 5% non-fat milk for 1 hour at room temperature and incubated overnight with anti-pRb (Cell signaling Technology; #8516, 1:1000) at 4 °C. Membranes were probed with fluorophore-conjugated anti-mouse or anti-rabbit secondary antibodies (1:10,000; ThermoFisher Scientific). Western blots were developed using the LI-COR Odyssey CLx imaging system (LI-COR Inc.) and quantitated using the Image Studio Lite software. The dose of ribociclib (0.5-5 uM) used per cell line was empirically determined as the dose at which phosphorylated Rb protein expression was reduced by 50 percent.

### Ribociclib-treated *in vitro* single cell RNA sequencing and analysis

The ribociclib and DMSO control experiments were performed in a passage-controlled fashion such that cells were split from the same parental line and then were allocated to either receive ribociclib or DMSO for a duration of 5 days. Following the fifth day of drug exposure, cells were harvested, cryopreserved to be processed at a later point, or immediately processed for single cell RNA sequencing (10x Genomics, 3’ version 3). All replicates for GB126 and replicates 1 and 3 GB86 targeted 6,000 cells for input into the 10x Chromium machine. Replicate 2 GB86 and all replicates for GB239 targeted 10,000 cells for input. Importantly, all matched replicates (e.g., replicate 1 for ribociclib and DMSO were processed under the same conditions). The single cell RNA sequencing data was then processed via cellranger count and Seurat. Briefly, cells were filtered out based on the following criteria: minimum number of genes detected > 500 and < 10,000 with a maximum percentage of mitochondrial genes set to 10%. These data were then processed by a Seurat workflow that normalization, highly variable gene detection, scaling, and Harmony batch correction [24] with the batch set to each experimental replicate group (e.g., GB239 replicate 1 for ribociclib and DMSO), followed by dimensionality reduction, UMAP visualization, and clustering. Large scale genetic events were inferred using inferCNV with the reference cells set to the myeloid and oligodendrocytes identified in the ribociclib-everolimus tumor sample dataset. Malignant cell state assignment was determined as described above for the ribociclib-everolimus-treated tumor samples. Statistical assessment was made between the malignant proportions of each cell state using a two-sided paired t-test for the experimental replicate.

### Statistical methods

Descriptive statistics of the clinical data are included Table 1. Patient characteristics were summarized using count and frequency for categorical variables, and median and interquartile range for continuous variables. Comparisons between groups were conducted using a two-sided or paired two-sided t-test, where indicated. Pearson’s correlation was used to investigate associations between ribociclib concentrations and cell type abundance. Multiple hypothesis test correction (i.e., false discovery rate correction) was applied for snRNAseq pseudobulk differential expression analysis. No data were excluded from the analyses, and no randomization was applied in correlative experiments. No adjustments were made for multiple comparisons and experiments and outcome assessment were not blinded. Data distribution was assumed to be normal unless otherwise indicated. P < 0.05 was considered significant. All statistical tests described were two-sided.

### Data and code availability

All analyses were performed in R 4.2.0. Gene expression count matrices processed by cellranger will be made available via the Gene Expression Omnibus (accession number available once deposited) and analysis code will be made available via github upon publication.

