## Supplementary Figures 1-10 for "Single nucleus transcriptomics, pharmacokinetics, and pharmacodynamics of combined CDK4/6 and mTOR inhibition in a phase 0/1 trial of recurrent high-grade glioma"

### Extended Data Figure 1

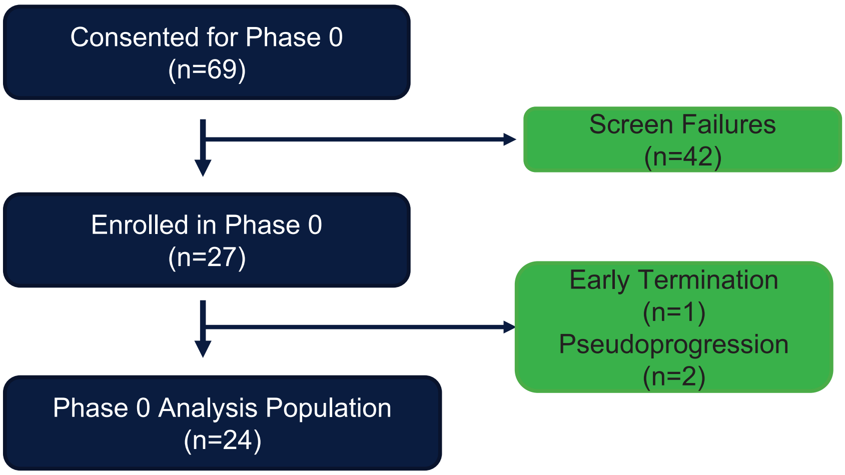


**Extended Data Fig. 1. CONSORT diagram.** Blue boxes on the left represent total number of patients consented, enrolled, and ultimately analyzed in this dataset. The green boxes present the number of patients that either failed to meet screening criteria or were not evaluated for pharmacokinetic and pharmacodynamic analysis due to early termination or treatment-associated pseudoprogression. Treatment-associated pseudoprogression was determined based on pathological findings from patients whose tumor was resected following drug exposure.

### Extended Data Figure 2

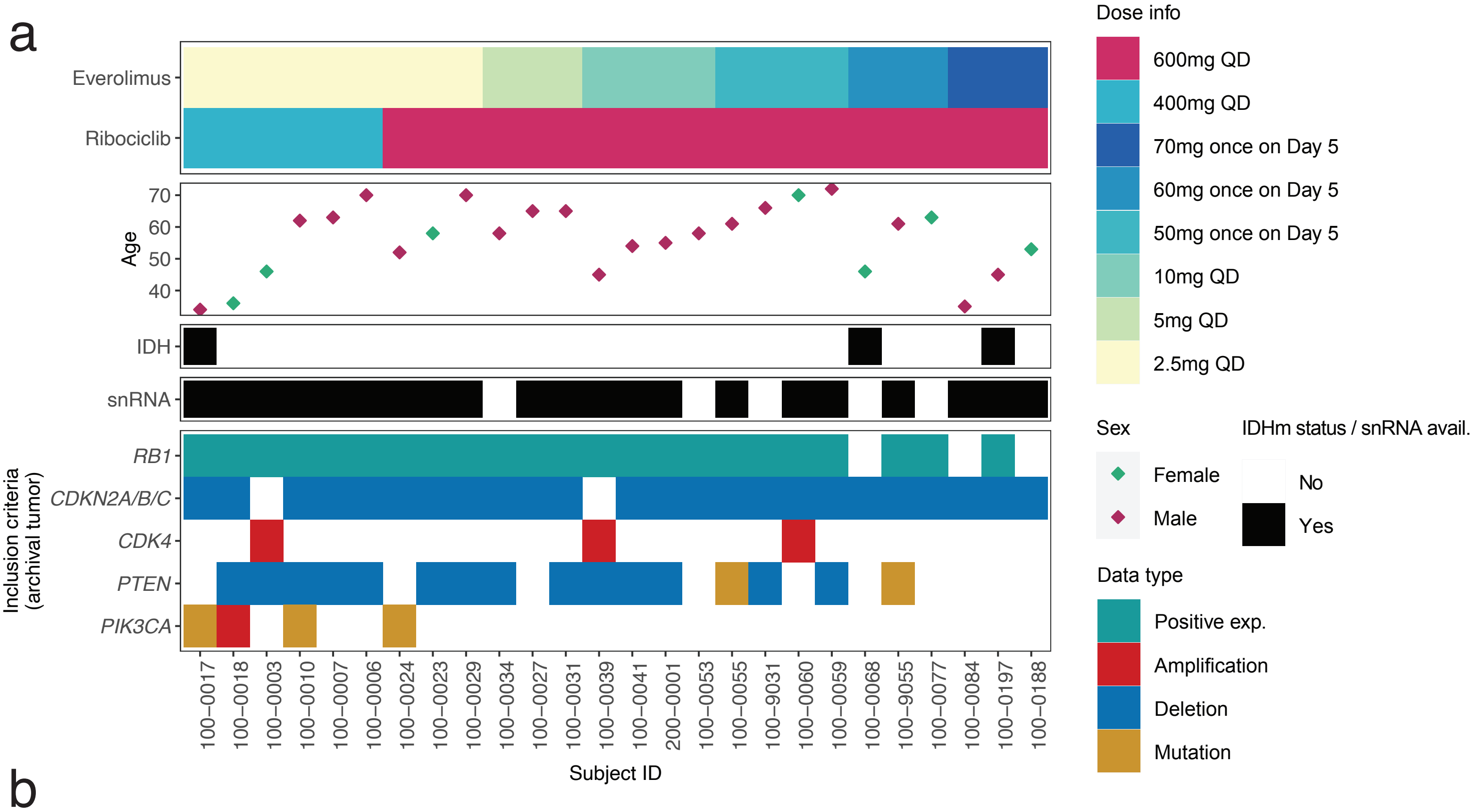


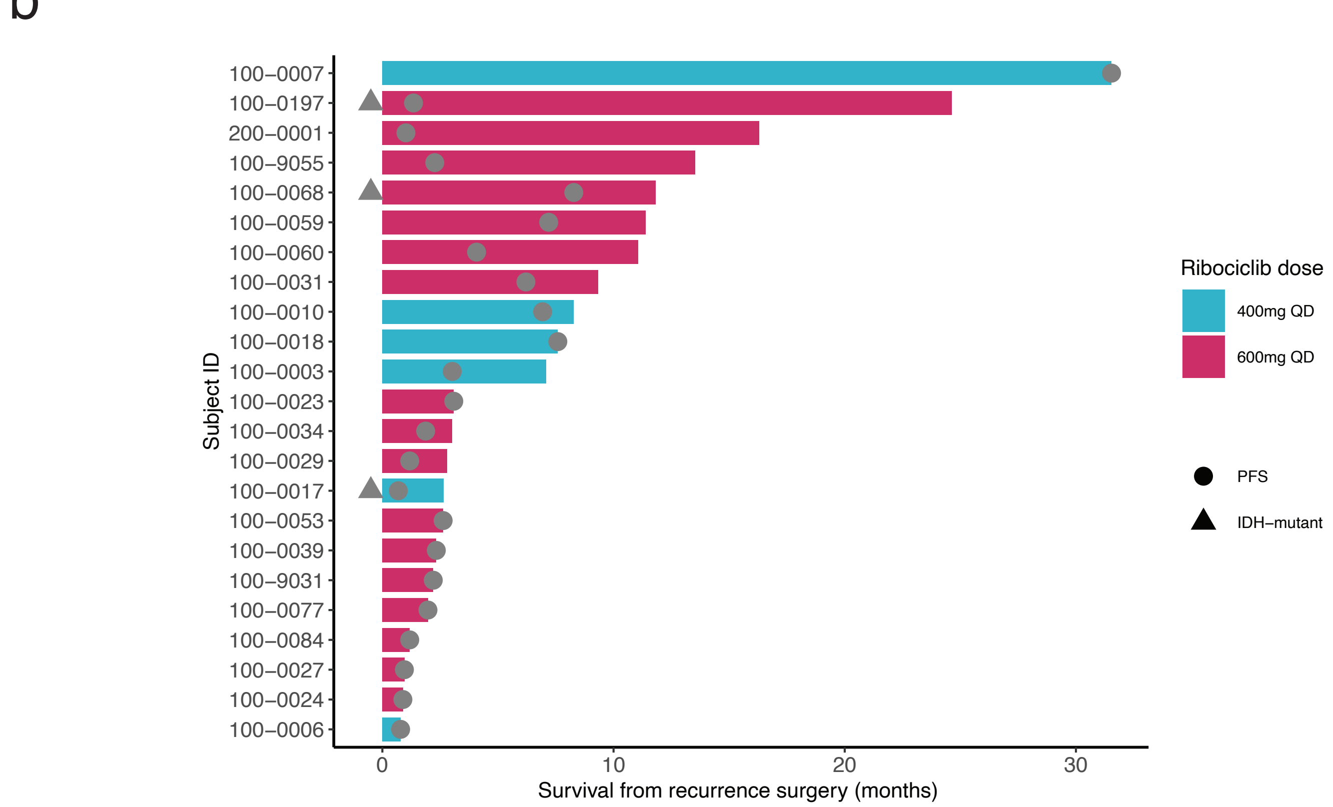


**Extended Data Fig. 2. Ribociclib-everolimus phase 0 clinical characteristics and survival outcomes.** (a) The x-axis reflects individual samples and study identifier. Each y-axis feature highlights a distinct tumor characteristic including (from top to bottom): dose of everolimus and ribociclib, subject age at tumor resection, subject sex, whether the archival tumor sample harbored an IDH mutation, whether snRNA data was generated for that tumor sample, Rb protein expression, and gene copy number and mutation information. (b) Patients for which survival information was available are presented as a swimmer plot. The color indicates the different ribociclib dose, the line represents death or last known follow up. Progression-free survival is noted as gray circle and IDH mutation status is indicated by a gray triangle.

### Extended Data Figure 3

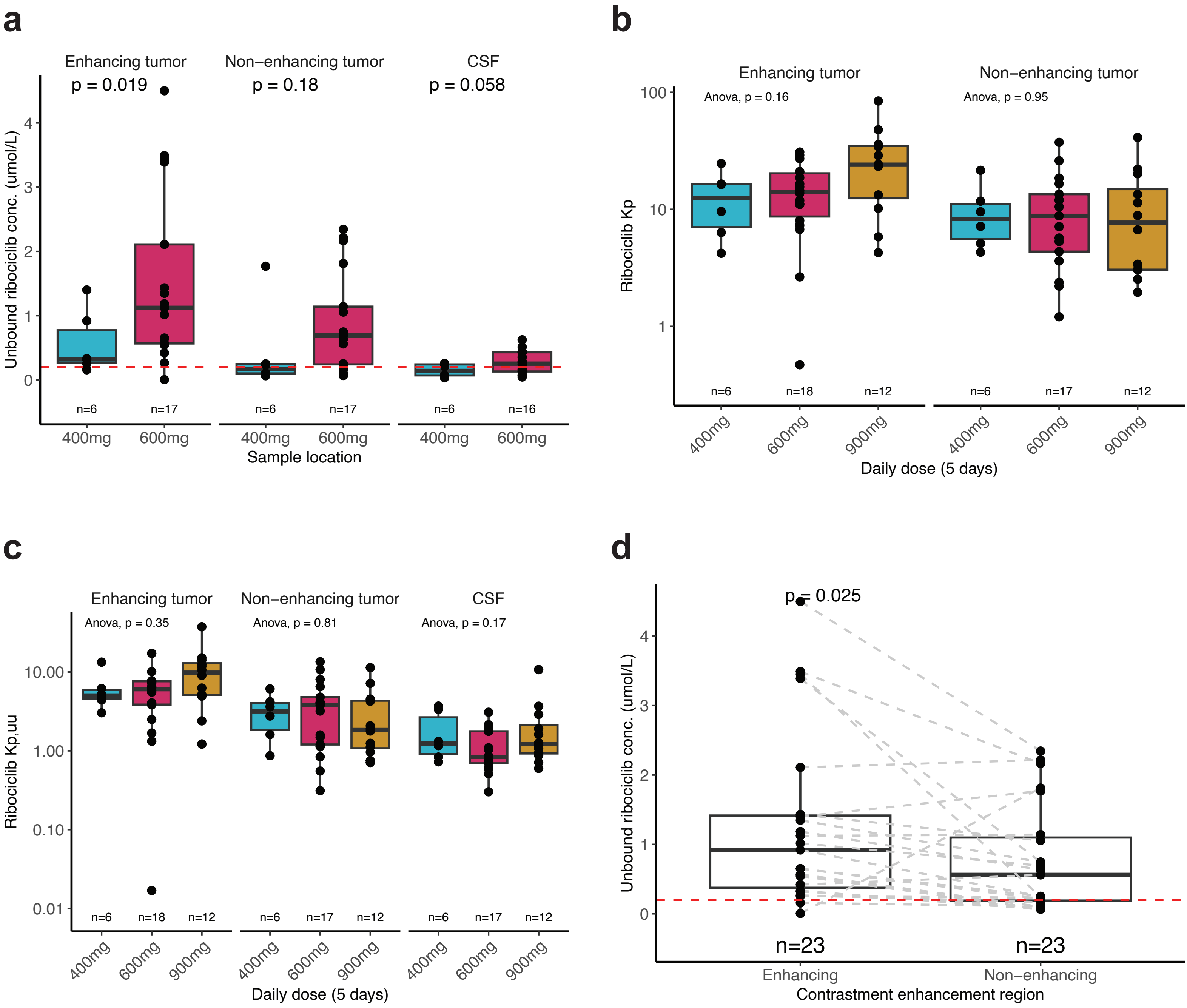


**Extended Data Fig. 3. CNS penetration of ribociclib in glioma patients following 5 days of treatment.** (a) Total ribociclib concentrations in the enhancing, non-enhancing, and plasma samples are presented for daily dose of 400 and 600 mg. Statistical differences were assessed with two-sided t-test. Boxplots represent the median, lower and upper quartiles, and the whiskers indicates 1.5 times the interquartile range. Each point represents an individual measurement. The dotted red line represents 5 times IC50 concentration for ribociclib (0.2umol/L= 5*0.04umol/L), which is a PK target in this current study. (b) Total drug tumor-to-plasma partition ratio (Kp). Each color represents a different dose, and each point a single measurement. The data for 900 mg were collected in a previous ribociclib monotherapy in high-grade glioma study and included for analysis here to aid dose comparisons [8]. (c) Unbound drug tumor- or CSF-to-plasma partition ratio (Kp,uu). Each color represents a different dose, and each point a single measurement. (d) Pairwise comparisons for unbound ribociclib concentrations comparing samples taken from the contrast enhancing and non-enhancing regions of a tumor. The data distribution is represented by a boxplot with each point indicating a sample. Dotted gray lines connect a patient's two values. The dotted red line represents 5 times IC50 concentration for ribociclib. The p-value for a paired two-sided t-test is shown.

### Extended Data Figure 4

**a**

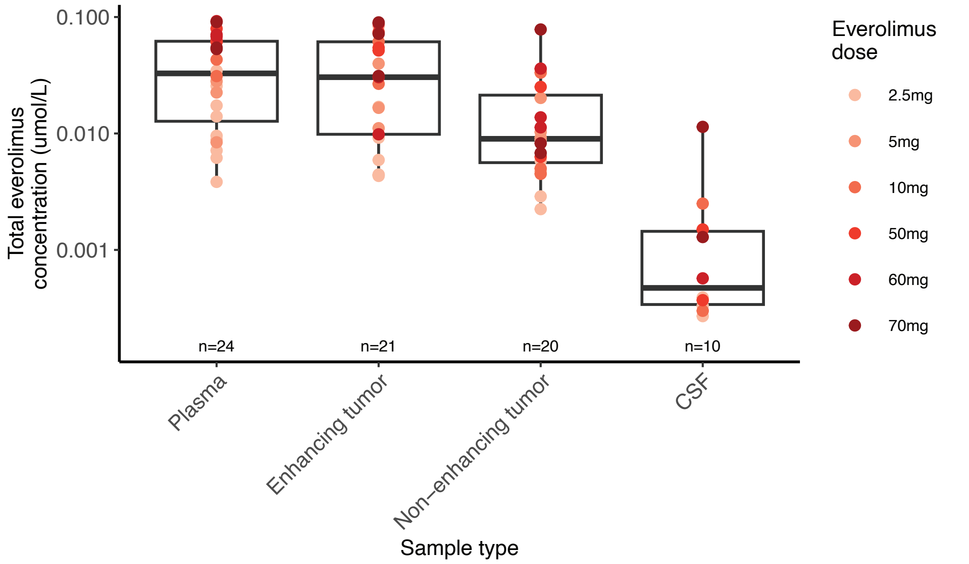


**b**

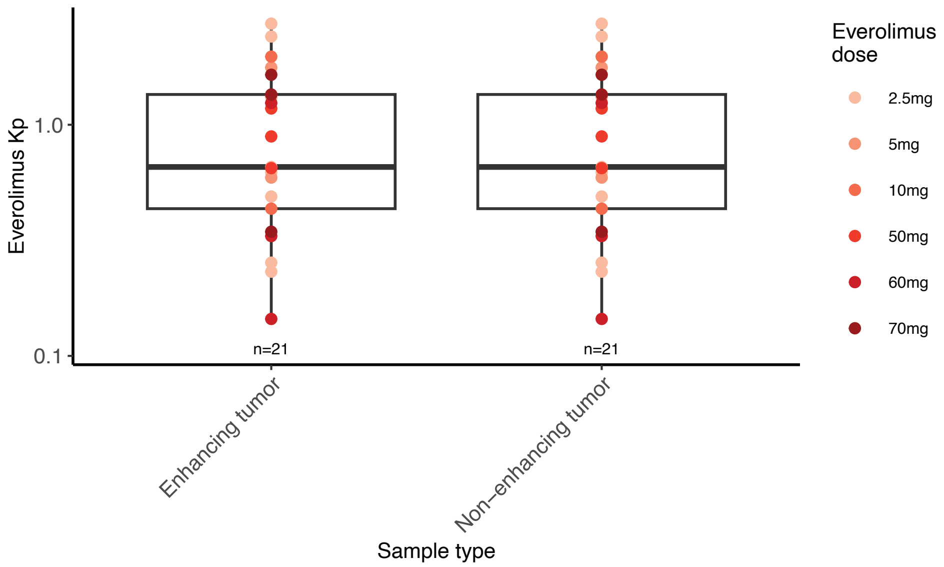


**Extended Data Fig. 4. CNS penetration of everolimus in glioblastoma patients following treatment.** Daily oral dose of 2.5, 5, and 10 mg, as well as a single oral dose of 50, 60, and 70 mg are presented. (a) Total drug concentrations in plasma, the enhancing and non-enhancing tumors, and CSF. Everolimus tumor concentration of 0.2 nM and CSF concentration of 0.1 nM represent below the lower limit of quantitation. Color represents everolimus dose. (b) Total drug tumor-to-plasma partition ratio (Kp). Kp of 0.001 represents below the lower limit of quantitation.with the median and range.

### Extended Data Figure 5

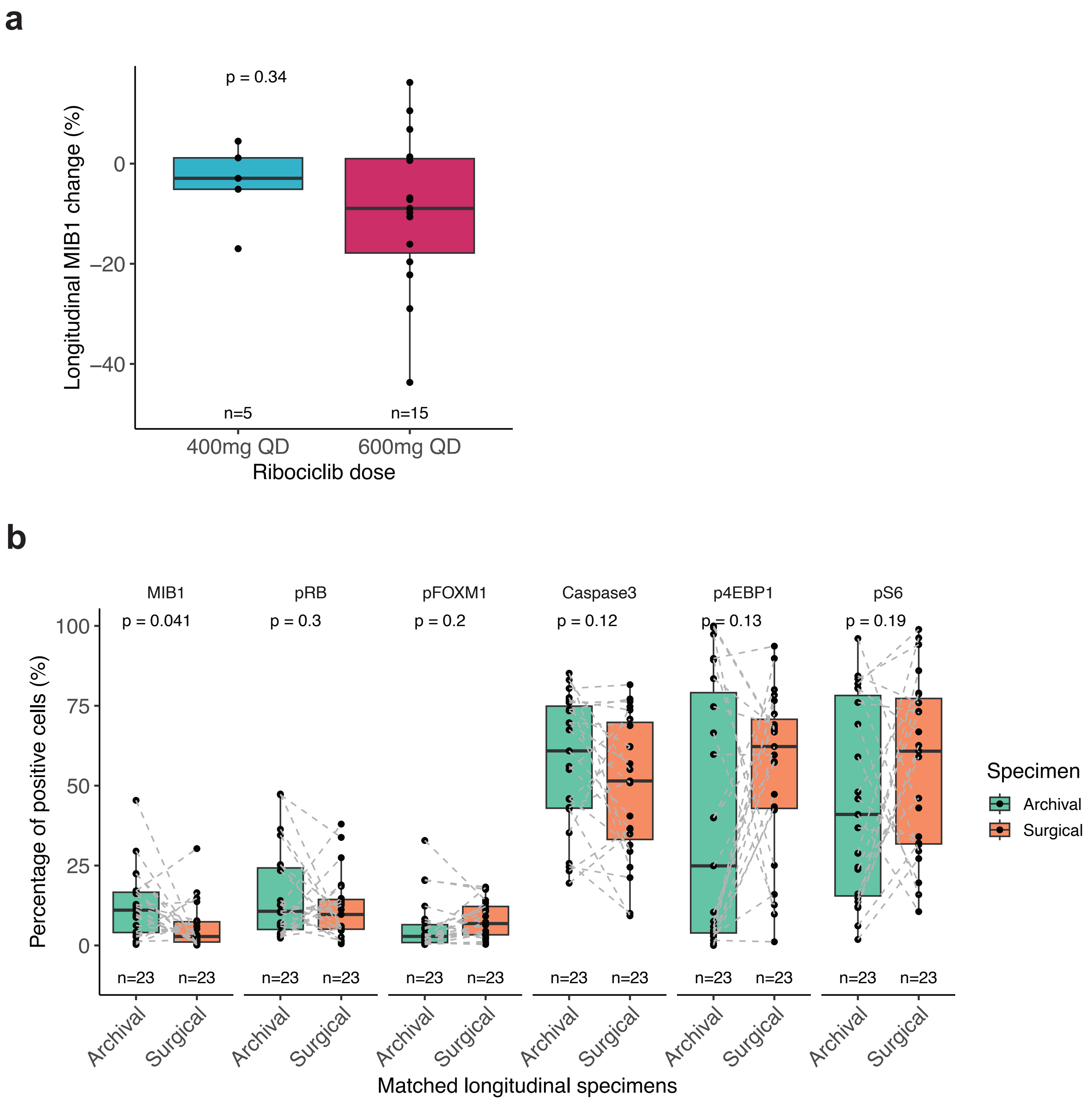


**Extended Data Fig. 5. Longitudinal changes in pharmacodynamic markers following targeted inhibition.** (a) Longitudinal difference in MIB1 (Ki67) proliferation marker separated by ribociclib dose (400mg and 600mg). P-value represents two-sided t-test. (b) All pharmacodynamic markers of ribociclib (MIB1, pRb, pFOXM1) and everolimus (Caspase 3, p4EBP1, and pS6) for both IDH-wildtype (n = 20) and IDH-mutant tumors (n = 3) are presented for archival and surgical tissues (i.e., post-ribociclib and everolimus) as box plots with individual points. Dotted gray lines connect a patient's two values.  P-values represent paired two-sided t-test.

### Extended Data Figure 6

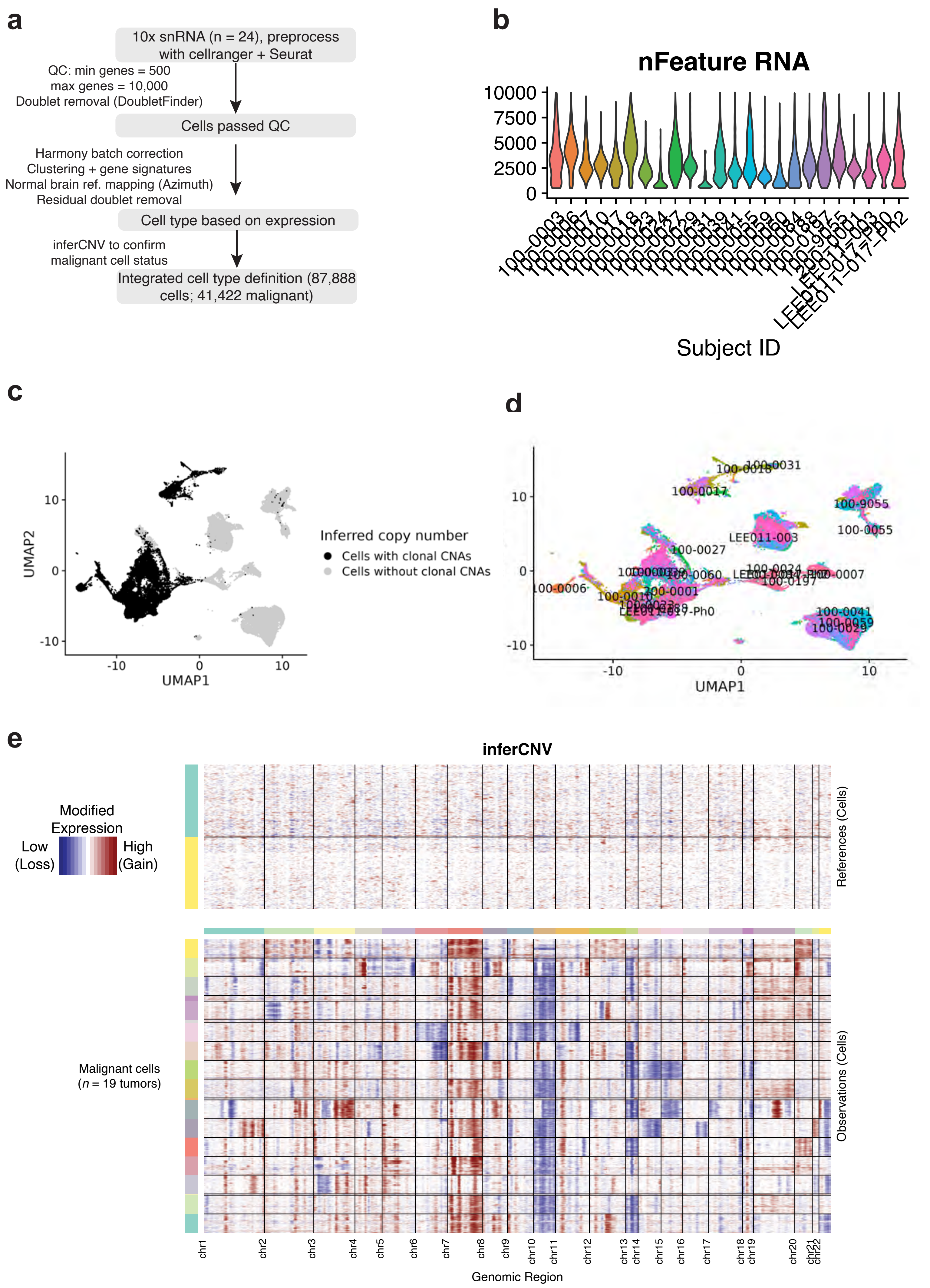

**Extended Data Fig. 6. Single nucleus data quality control and cell type identification.** (a) workflow for quality control filtering and cell type identification in phase 0 human glioblastoma samples (b) Number of genes detected for each sample with snRNA data (n = 24 samples) (c-d) The same UMAP image presented in Fig. 3 colored by (c) inferred copy number alterations that have previously been described as clonal [4, 5, 24] (i.e., occurs early in tumor development) high-grade events (Chr7 amplification and Chr10 deletion) and (d) subject identifier. (e) Cohort-level inferred copy number alteration visualization. Chromosomal location is presented along the x-axis with each line separating chromosomes. Y-axis colors reflect individual samples from phase 0 study. The top-panel represents reference cells used to infer copy number status (i.e., myeloid cells and oligodendrocytes). While copy number alteration values were determined for all cells, samples with more than 500 malignant cells were down sampled to ensure more even distribution of cells across samples. Heatmap color reflects copy number gains (red) and losses (blue).

### Extended Data Figure 7

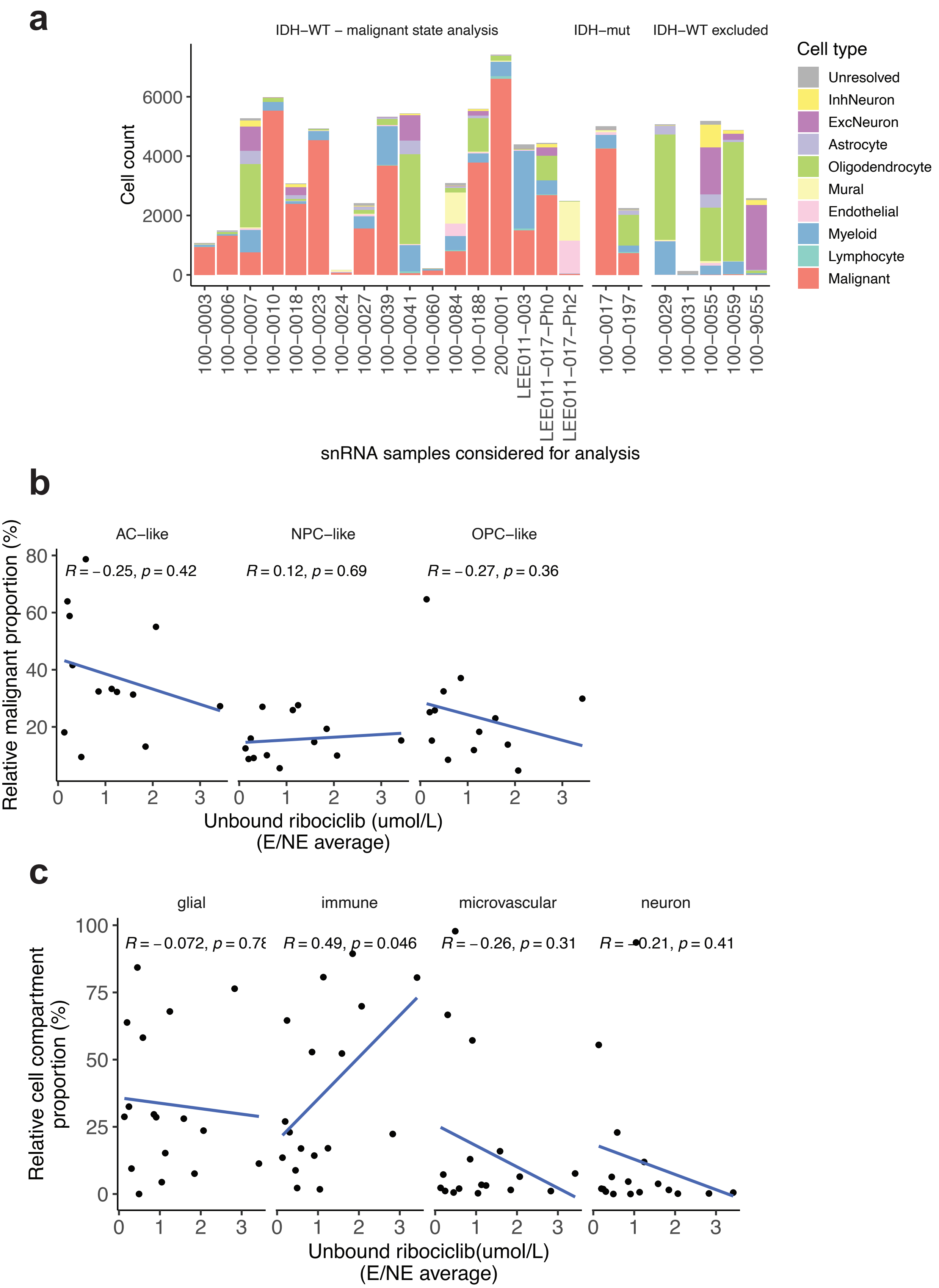


**Extended Data Fig. 7. Cell type proportion and associations with drug response.** (a) Stacked bar plots represent the total number of cells per sample, colored by the cell type, and faceted by analysis type. IDH-wild-type samples with insufficient malignant cell abundance were removed from malignant state analyses. (b) The correlation between the unbound ribociclib concentration average across the enhancing and non-enhancing regions of each tumor are plotted against the relative malignant cell proportions for each malignant state that were not shown in Fig. 3e. The Pearson correlation coefficient and p-value is shown for each cell type. (c) The correlation between the unbound ribociclib concentration average across the enhancing and non-enhancing regions of each tumor are plotted against the relative tumor microenvironment proportions. The Pearson correlation coefficient and p-value is shown for each tumor microenvironment compartment.

### Extended Data Figure 8

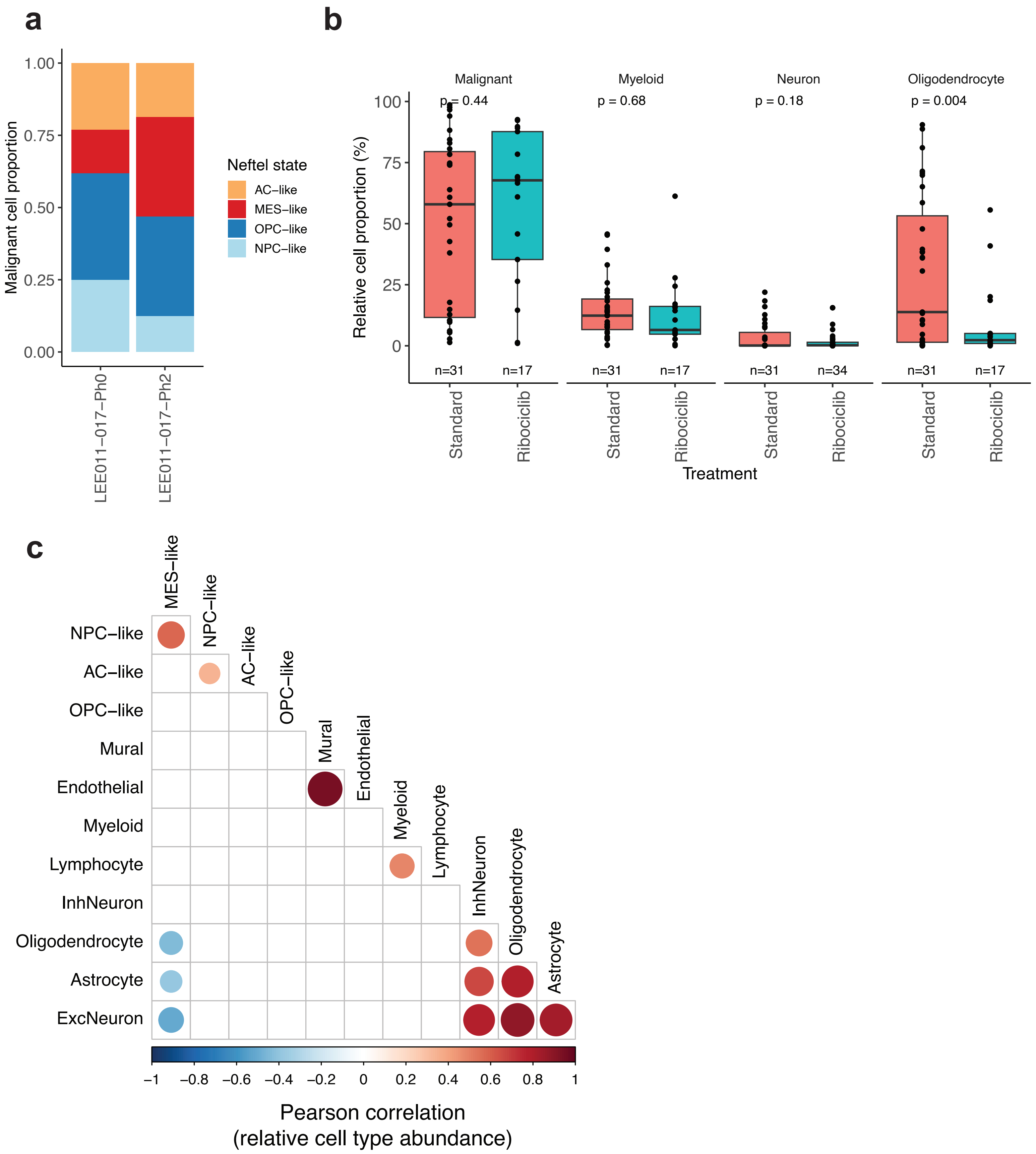


**Extended Data Fig. 8. Longitudinal and treatment associated changes in ribociclib-treated tumors.** (a) The longitudinal difference in cell type proportions for single tumor that was treated with ribociclib monotherapy in phase 0 and phase 2. Presented are the malignant cell states (n = 2,668 phase 0 and n = 33 phase 2 malignant cells). (b) The box plots represent the relative proportion of tumor microenvironment cell types for both the standard treatment and post-ribociclib treatment groups for the five most abundant cell types. (c) Heatmap representing the Pearson correlation values between cell state abundance (both malignant and non-malignant) in ribociclib-treated samples. Dot color and size represents strength of association. Only correlations where adjusted p-value was less than 0.05 are shown. All non-significant correlations are shown as a blank white tile.

### Extended Data Figure 9

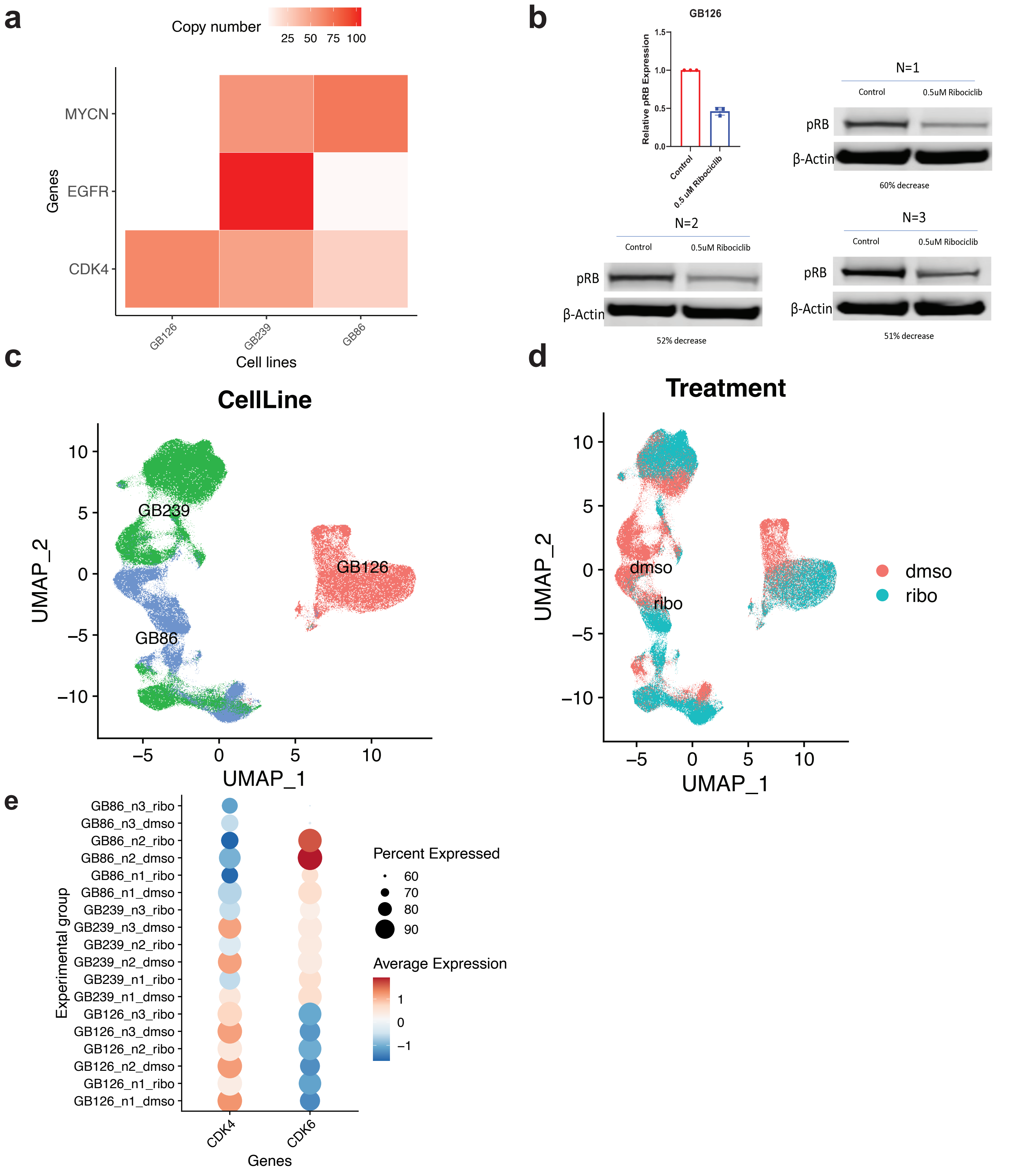


**Extended Data Fig. 9. In vitro ribociclib perturbation and response.** (a) Panel sequencing of 3 patient-derived IDH-wild-type glioblastoma cell lines that harbor CDK4 and other oncogene amplifications. Increasing color intensity represents estimated copy number for cell lines. CDK4 amplification was the only high-level amplification detected in GB126. (b) Representative results from dose-determination that inhibited pRb levels by 50%. Shown are three replicates for the 0.5uM dose that resulted in consistent decrease and was selected for in vitro single-cell RNA sequencing experiments for GB126. (c-d) UMAP representation of single cell data indicate both cell line and treatment-associated differences. (e) Gene expression dot plots indicate the relative expression levels of CDK4 and CDK6 (color) as well as the percentage of cells within a sample that are expressing the indicated gene. The y-axis presents each cell line, the passage controlled experimental replicate (e.g., “n1”), and whether the sample was treated with ribociclib (ribo) or DMSO.

### Extended Data Figure 10

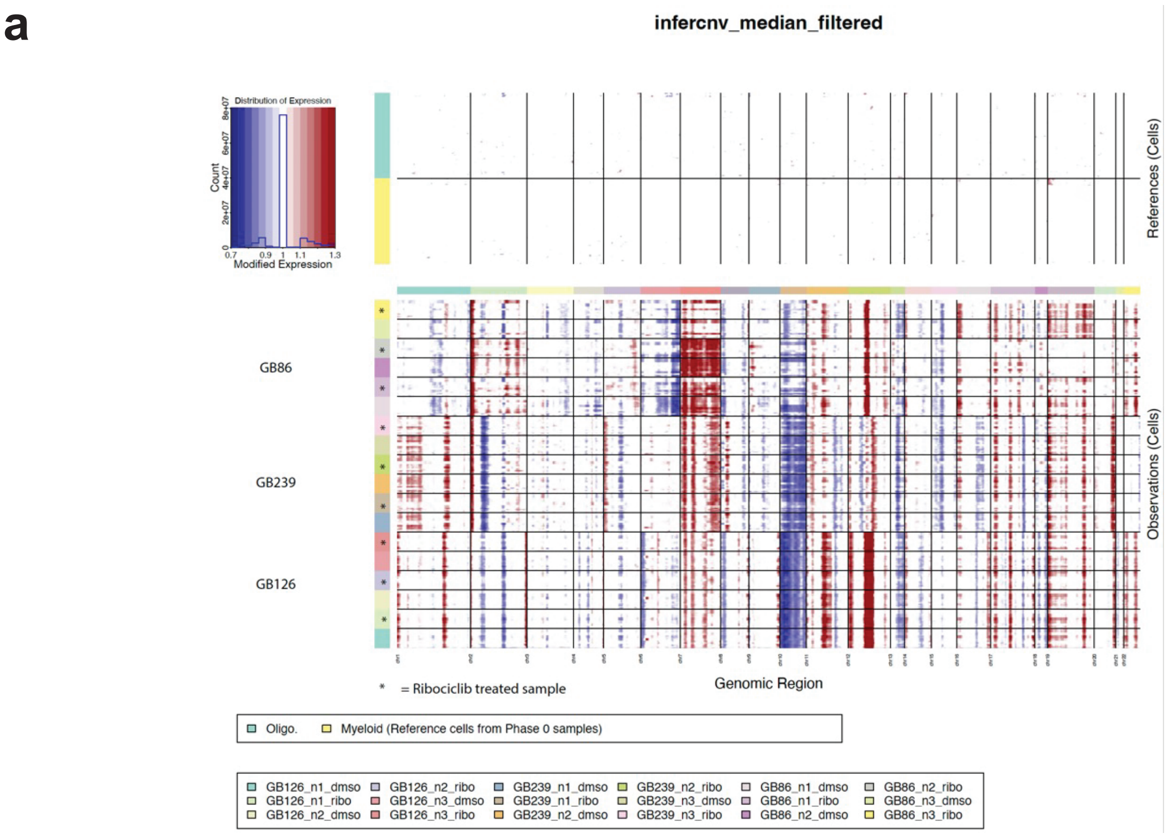


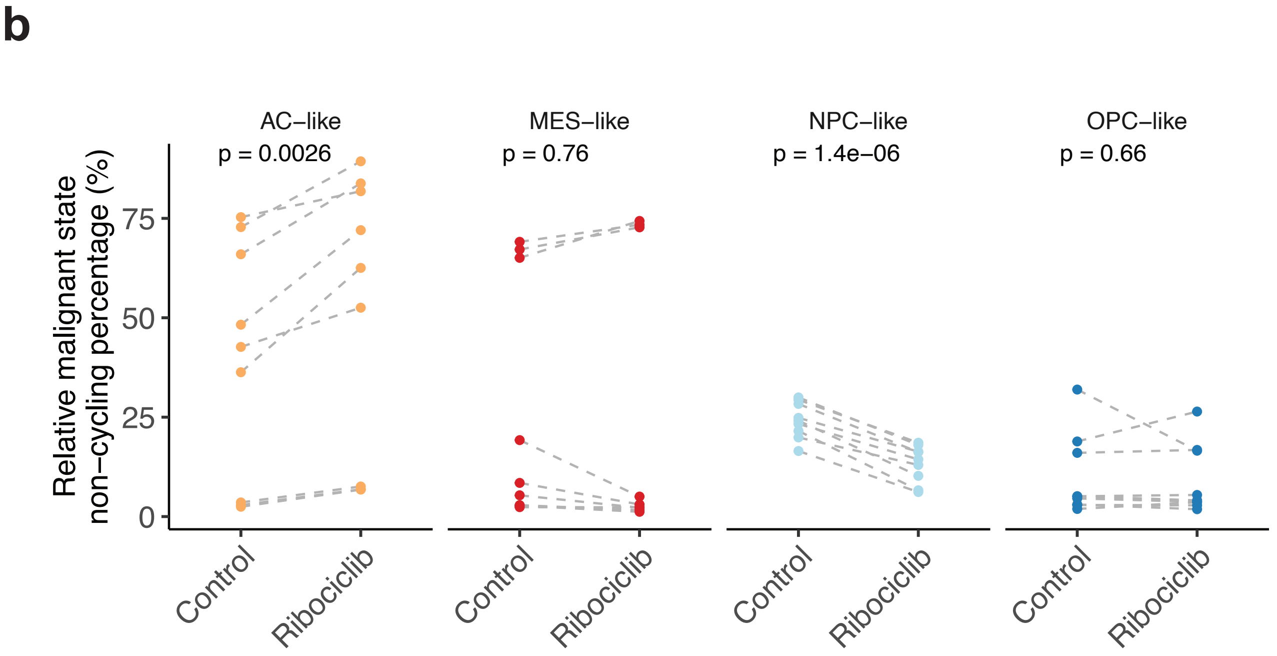


**Extended Data Fig. 10. In vitro inferred genetic stability and malignant cell state shift following ribociclib exposure.** (a) Inferred copy number alterations for in vitro ribociclib versus DMSO analyses. The top panel reflects reference cells, myeloid cells, and oligodendrocytes, from the phase 0 tumor samples. The bottom panel reflects the cell line inferred copy number amplifications (red) and deletions (blue). The x-axis represents sequential ordering of chromosomal location. The y-axis represents each individual sample as indicated by the color block for each cell line. The y-axis is ordered such that each experimental replicate of ribociclib (indicated by a "*") and DMSO are shown vertically next to one another. (b) Non-cycling malignant cell state proportions are shown for both the DMSO control and ribociclib exposed cell lines. The plot is separated by cell state and all replicates across cell lines are presented together. The gray dotted line within each panel denotes an experimental replicate that connects the DMSO and ribociclib for that group. The p-value represents a two-sided paired t-test.
